# Mortality Attributable to Drought, Wildfire Smoke, and Their Concurrent Added Effects in the Contiguous United States

**DOI:** 10.1101/2025.09.24.25336547

**Authors:** Ying Hu, Lingzhi Chu, Pin Wang, Azar M. Abadi, Minghao Qiu, Kai Chen

## Abstract

Climate change has led to more frequent individual and concurrent drought and wildfire smoke events in the U.S., yet whether their concurrence adds to health burden remains understudied. We assessed the frequency of these events, developed a Two-Step Individual and Added Effect Estimation (TIAE) Model to evaluate their effects, and estimated the attributable mortality across 3,103 U.S. counties from 2007 to 2023. We identified annual averages of 3,630 county-months (∼110,342 county-days) of drought, 14,049 county-days of wildfire smoke, and 980 county-days of concurrent exposure. Both individual events were significantly associated with increased all-cause mortality, with a significant added effect observed during concurrent days. We estimated 6,576 (95% CI: 3,990, 9,155), 10,465 (95% CI: 6,642, 14,261), and 469 (95% CI: 256, 682) annual deaths attributable to drought, wildfire smoke, and their added effect, respectively, with a higher burden in counties with higher overall social vulnerability index. These findings call for targeted measures to address the burden from drought, wildfire smoke, and their concurrence.

## Introduction

Droughts are becoming increasingly intense and persistent in many regions worldwide^1^. These trends are driven by climate change, through rising temperatures, altered precipitation patterns, and disruptions of the global hydrological cycle, such as the El Niño Southern Oscillation^2,3^. In addition, long-term declines in soil moisture and human behavior influence further exacerbate drought conditions^4,5^. Drought also increases wildfire risks through influencing multiple factors related to fire ignition, spread, and regime across various scales^6^. Reduced fuel moisture during droughts, a key determinant of fire weather, has been shown to strongly correlate with interannual variability in burned area in the western U.S.^7-9^. Extended drought can further alter vegetation species composition, structural patterns, and landscape characteristics, influencing regional fire regimes^10^. As climate change accelerates, rising temperatures and other shared drivers are expected to increase the frequency of both droughts and wildfires in the U.S.^2,11^, and might also increase their co-occurrence. Understanding the health impacts of these events is therefore essential for informing effective public health and climate adaptation strategies.

A comprehensive understanding of the health burden of compound drought-wildfire smoke events requires not only characterizing the individual health impacts of drought and wildfire smoke but also exploring whether their concurrence poses an additional burden. Previous studies have shown significant association between drought and increased mortality in the U.S., along with evidence of differences in population vulnerability. Berman et al. ^12^ and Gwon et al. ^13^ found elevated risks of cardiovascular diseases (CVD) and all-cause mortality during drought periods in some regions in the U.S. Lynch et al. ^14^ found that the mortality impacts of drought may vary geographically, while Abadi et al. ^15^ observed a greater association among middle-aged white populations in Nebraska. However, the national mortality burden attributable to drought in the U.S. remains unclear. Wildfire smoke exposure, particularly from fine particulate matter (particulate matter with an aerodynamic diameter less than 2.5 μm, PM_2.5_) ^16^, is increasingly recognized to elevate risks of premature death from multiply causes, such as CVD, respiratory diseases, endocrine, nutritional, and metabolic diseases, digestive diseases, and mental health-related deaths^17,18^. Recent estimates suggest that wildfire smoke contributes to thousands to tens of thousands of premature deaths annually in the U.S.^19-23^. Evidence on whether concurrent drought and wildfire events pose an added health burden remains limited. One study in low- and middle-income countries found wildfire smoke may mediate drought effects on child growth^24^, while another reported that compound drought and heatwave events tend to increase mortality risk among patients with COPD^25^. These findings imply that drought could amplify the impacts of other extreme events, yet they do not directly address whether concurrence with wildfire smoke entails an added effect. Overall, it remains unclear how many deaths could be attributable to drought individually, or whether an added burden arises when drought and wildfire smoke occur concurrently. These important knowledge gaps hinder our understanding of the comprehensive health impacts of drought and wildfire smoke events and limit our ability to identify vulnerable populations.

Analyzing the added effect of concurrent exposure to drought and wildfire smoke poses methodological challenges. Previous studies have commonly employed the Relative Excess Risk Due to Interaction (RERI), Attributable Proportion Due to Interaction, or Synergy Index to quantify added association of concurrent extreme events, typically by comparing health risk on days with concurrent exposure to those without^26-29^. However, these approaches require binary individual and concurrent exposures assessed at the same time window and have been mostly applied to study short-term (e.g., daily) exposures. Extending such methods to assess the health impacts of longer-term concurrent exposures (e.g., monthly or annually) to multiple events is challenging. For example, individuals often experience mixtures of individual and concurrent exposures within a year, making it hard to differentiate individual and concurrent exposures using a binary exposure variable at the annual time window. Moreover, individual and concurrent exposures, when included simultaneously in statistical models, could lead to biased or unrobust association estimates due to their potential collinearity. Addressing these limitations requires modeling approaches that can disentangle individual effects and added effects from concurrent exposures while mitigating collinearity when evaluating association between long-term exposures to multiple extreme events and health outcomes.

This study aims to provide a comprehensive understanding of drought, wildfire smoke and their concurrence in the U.S., from their exposure patterns, individual and concurrent added health effect, and associated mortality burdens. We first summarized the annual average number of drought months, wildfire smoke days, and their concurrent days across 3,103 U.S. counties from 2006 to 2023, as daily wildfire smoke PM_2.5_ are available for these years. We then developed a Two-Step Individual and Added Effect Estimation (TIAE) model that isolates the individual and concurrent added associations through a two-step analysis, minimizing potential confounding between them (**Figure S1**). We utilized this model to estimate both the individual effect of drought and wildfire smoke exposure, as well as the added effects of their concurrence over the current and preceding 11 months on cause-specific mortality, thereby capturing long-term exposure impacts. Finally, we estimated the annual number of deaths attributable to drought, wildfire smoke, and the additional deaths attributable to their concurrence in the contiguous U.S. from 2007 to 2023. We also conducted stratified analyses by sex, age, and counties in different Social Vulnerability Index (SVI) groups, aiming to identify vulnerable populations.

## Results

### Exposure Patterns of Drought, Smoke, and Their Concurrent Events

Standardized Precipitation Evapotranspiration Index (SPEI) is a multiscale drought index that simultaneously accounts for both climatic water supply (precipitation) and atmospheric water demand (potential evapotranspiration). It quantifies how anomalously dry or wet a given period is compared to historical norms, with negative values (SPEI < 0) indicating drier-than-normal conditions (i.e., drought), and positive values (SPEI > 0) indicating wetter-than-normal conditions^30^. Because it can be calculated over different time scales (e.g., 1-month, 3-month, 6-month, 12-month), SPEI is particularly useful for assessing both short-term and long-term hydroclimatic variability and its potential impacts on health, agriculture, and ecosystems. In this study, we defined a drought month as a month with a population-weighted 1-month SPEI below −1.2, approximately corresponding to the 10^th^ percentile county-level monthly SPEI during the studied period. We selected the 1-month SPEI to capture short-term meteorological drought conditions. The 10^th^ percentile has previously been used to identify drought based on SPEI and Standardized Precipitation Index (SPI) ^31,32^. We defined a wildfire smoke day as a day with daily wildfire smoke PM_2.5_ concentrations reached or exceeded 10 µg/m^3^, approximately corresponding to the 99^th^ percentile during the studied period. Concurrent drought and wildfire smoke days were identified as wildfire smoke days within a drought month. Between 2006 and 2023, across 3,103 U.S. counties, there was an annual average of 3,630 county-months of drought (9.7% of all county-months) and 14,049 county-days of wildfire smoke (1.2% of all county-days), with an annual 980 county-days when both occurred (0.08% of all county-days). At the county level, the mean annual exposure was 1.2 months for drought (min–max: 0–2.1), 4.5 days for wildfire smoke (min–max: 0.2–23.9), and 0.3 days for their concurrent events (min–max: 0–2.1). We also explored exposures under alternative SPEI timescales (3, 6, and 12 months) and thresholds (−1.0, −1.1, −1.3, −1.4, and −1.5), as well as daily wildfire smoke PM_2.5_ thresholds (5 and 15 µg/m^3^). Exposure days or months under these exposure metrics are provided in **Table S1**.

Analyzing temporal trends and spatial distributions over four periods (2006–2010, 2011–2015, 2016–2020, and 2021–2023, **Figure 1**), we observed clear shifts in both the intensity and geographic extent of exposure (census bureau-designated regions and divisions of the U.S. were shown in **Figure S2**). During 2006–2010, there was an annual average of 4,060 county-months of drought, mainly concentrated in the southern U.S. In 2011–2015, drought exposure rose to 4,238 county-months annually, with increased occurrence observed in parts of California, Nevada, Texas and extending northward into New Mexico, Oklahoma, Kansas, and Missouri, and surrounding areas. In 2016–2020, both drought intensity and spatial coverage declined to 2,680 county-months per year, primarily affecting Oregon and Washington. By 2021–2023, drought exposure rose again to 3,481 county-months annually, extending from Oregon and Washington into broader areas of the Pacific and northern Mountain regions.

**Figure 1.**
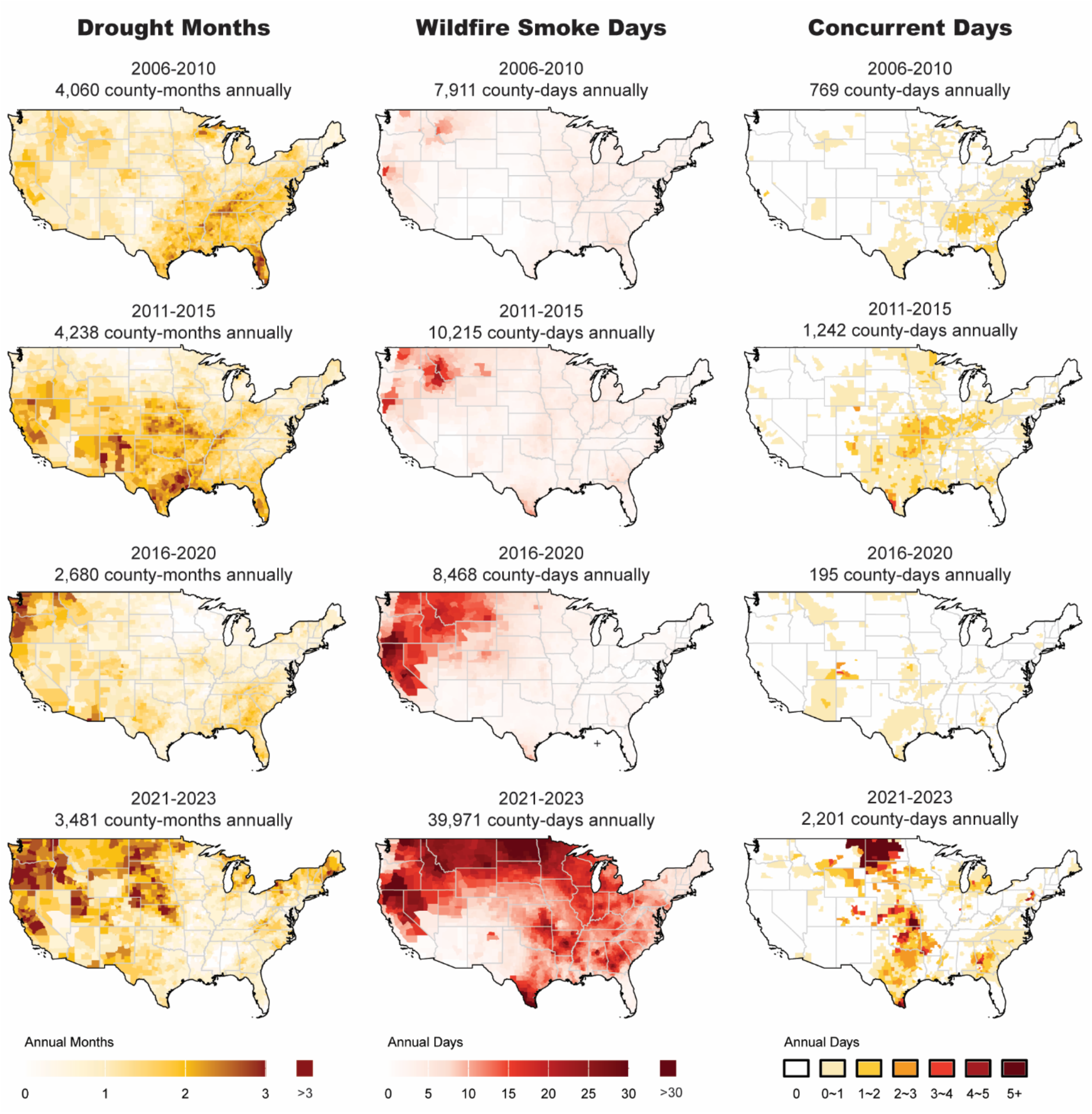
Spatial distribution of drought months, wildfire smoke days and their co-occurrence days across 3,103 U.S. counties over four periods (2006–2010, 2011–2015, 2016–2020, and 2021–2023).

For wildfire smoke, the annual average was 7,911 county-days during 2006–2010, with slightly higher exposure in parts of the Northwestern states. This rose to 10,215 county-days in 2011–2015, with most counties experiencing increased exposure, particularly in Montana, Idaho, Washington, and around the Oregon-California border. Although the annual average decreased to 8,468 county-days in 2016–2020, the impacted regions became more geographically concentrated. Exposure declined across the eastern U.S., while both the extent and frequency of smoke days increased in the Northwest, with a particularly notable rise in parts of California, Oregon, Montana, Idaho, Washington, and Nevada. By 2021–2023, wildfire smoke exposure surged to 39,971 county-days, with substantial increases across most of the country except for the southern Mountain region.

Spatial patterns of concurrent drought and wildfire smoke exposure more closely align with those of drought than with wildfire smoke. During 2006–2010, the annual average of concurrent was 769 county-days, mainly concentrated in the southern U.S. It increased to 1,242 county-days in 2011–2015, with affected areas extending across Texas and neighboring states, as well as parts of the Midwest. In 2016–2020, concurrent exposure declined to an annual average of 195 county-days, with notable decreases across most regions. By 2021–2023, concurrent exposure rose markedly to 2,201 county-days, with the severe impacts observed in the West North Central and West South Central regions, particularly in North Dakota.

We group counties into low (bottom 50%, 1551 counties) and high (top 50%, 1552 counties) SVI groups to explore exposure differences (**Figure 2**). All counties experienced at least one day of wildfire smoke exposure during the study period, and more than 99.9% experienced at least one month of drought. Most counties also experienced concurrent drought and wildfire smoke days, with a higher proportion in the high SVI group (88%) compared to the low SVI group (84%). A higher proportion of high SVI counties fell into the upper range of annual drought exposure (≥1.5 months per year), whereas low SVI counties were more concentrated in the lower range (<1 month per year) than high SVI counties. Wildfire smoke exposure was generally similar between the two groups, although the low SVI group showed slightly higher proportions exceeding 5 days per year. For concurrent drought and wildfire smoke exposure, the high SVI group exhibited marginally higher proportions in the higher range (≥0.5 days per year).

**Figure 2.**
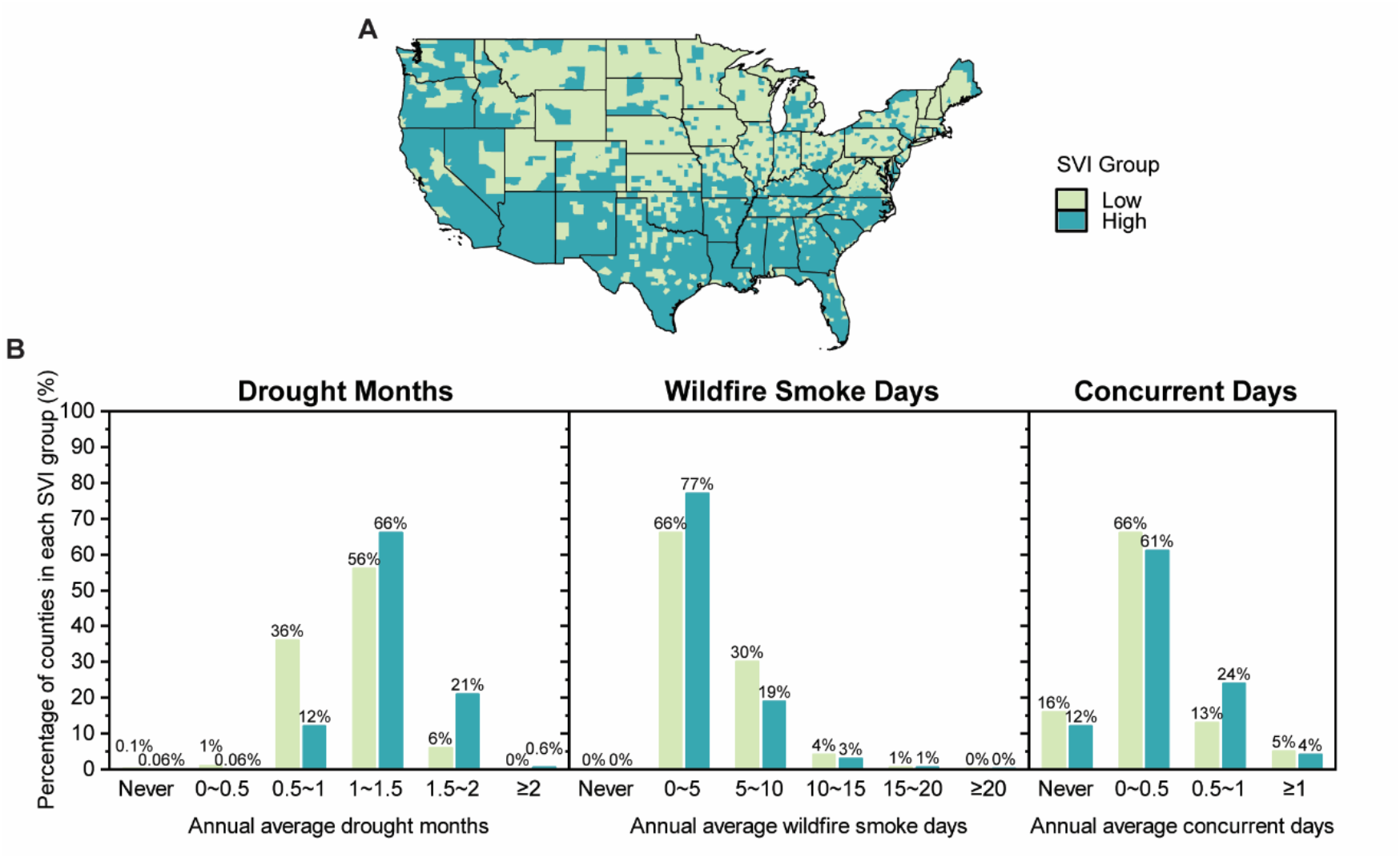
Annual average drought months, wildfire smoke days, and concurrent days (2006–2023) across 3,103 U.S. counties, by Social Vulnerability Index (SVI) group. A. SVI map. B. Event days in High (n = 1552). and Low SVI groups (n = 1551).

### Individual and Added Health Effects of Drought and Wildfire Smoke

Using a TIAE model developed in this study, we estimated the individual effects of drought month and wildfire smoke day exposure, as well as the added effect of their concurrence, on all-cause and cause-specific mortality. We focused on causes previous found to be associated with drought or wildfire smoke, including non-external, CVD, respiratory diseases, endocrine, nutritional, and metabolic diseases, digestive diseases, and mental and behavioral disorders deaths ^17,18^ (**Figure 3 and Table S2**). Details of the modeling approach are provided in the **Methods section**. We found increases in drought month and wildfire smoke day were both significantly associated with elevated monthly all-cause mortality, and their concurrence demonstrated a significant added effect. The relative risk (RR) associated with each additional drought month over the current and preceding 11 months and monthly all-cause mortality was 1.0029 [95% confidence interval (CI): 1.0018, 1.0041]. The RR for each additional wildfire smoke day during the same period was 1.0013 (95% CI: 1.0008, 1.0018), and the RR for the added effect of an additional concurrent day was 1.0009 (95% CI: 1.0005, 1.0013).

**Figure 3.**
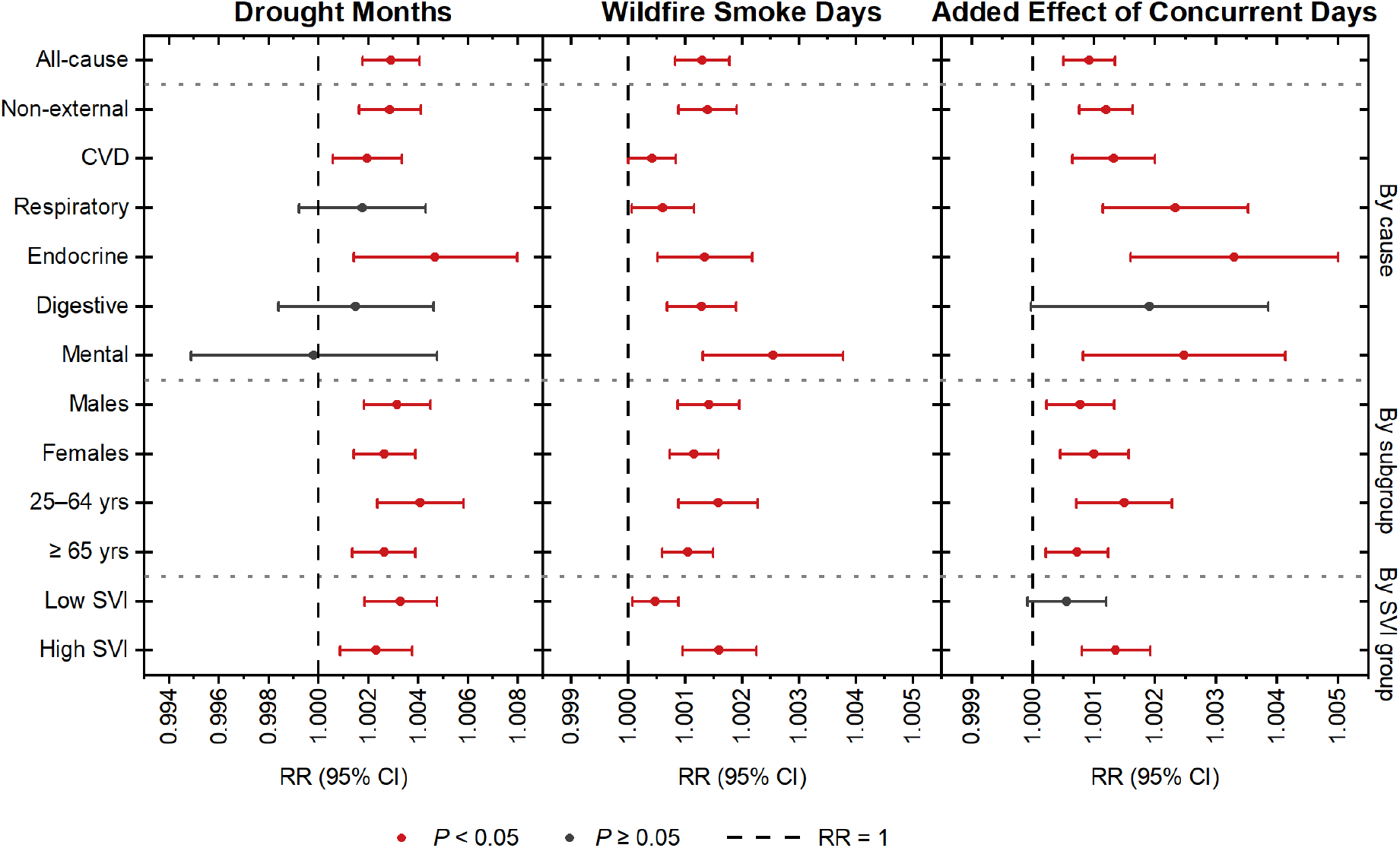
Relative risk (RR and its 95% CI) of monthly cause-specific mortality associated with per additional month of drought, per additional day of wildfire smoke, and per addition day of their concurrent during current and previous 11 months, as well as all-cause mortality for individuals from different subgroups. CVD: cardiovascular diseases; Endocrine: endocrine, nutritional and metabolic diseases; Mental: mental and behavioral disorders.

Our cause-specific analyzes also illustrated that an increase in drought months was significantly associated with monthly deaths from non-external causes, specifically CVD and endocrine, nutritional, and metabolic diseases, while no significant associations were observed for respiratory diseases, digestive diseases, or mental and behavioral disorders. An increase in wildfire smoke days over the same period was significantly associated with all cause-specific monthly mortality outcomes analyzed, including non-external, CVD, respiratory diseases, endocrine, nutritional, and metabolic diseases, digestive diseases and mental and behavioral disorders deaths. An increase in concurrent days of exposure was associated with a significant added risk in monthly mortality for all outcomes analyzed except digestive diseases, for which the association was of borderline significance. Notably, the added effect on CVD, respiratory, and endocrine mortality exceeded the corresponding effects observed for wildfire smoke days alone, while the impacts on non-external and mental and behavioral mortality were of similar magnitude.

Our subgroup analyses showed that for both sexes (male and female) and age groups (25–64 and ≥65 years), increases in drought months and wildfire smoke days over the current and preceding 11 months were each significantly associated with higher monthly all-cause mortality, with a significant added effect also observed for concurrent exposure days. However, we found no significant differences between these subgroups in association between events and all-cause mortality. Our analysis by SVI group revealed that increases in drought months and wildfire smoke days were associated with higher monthly mortality in both groups, but the added effect was only significant in high SVI counties. The impact of increased drought months did not differ significantly between SVI groups [RR difference = 0.9990, 95% CI: 0.9970, 1.0011], while the effect of wildfire smoke days was significantly higher in high SVI counties [RR difference = 1.0011, 95% CI: 1.0004, 1.0019] and the added effect of concurrent exposure was broadline significantly higher in high SVI counties [RR difference = 1.0008, 95% CI: 0.9999, 1.0017]. These differences suggest that populations with higher social vulnerability may face a disproportionate burden from compound drought and wildfire smoke events. RRs and RR differences of subgroup analyses by sex, age, and SVI for various cause-specific mortality outcomes are summarized in **Table S2**.

### Mortality Attributable to Drought, Wildfire Smoke, and Their Concurrent-Day Added burden

Because we focused on the exposure over current and the preceding 11 months as the long-term exposure, the year 2006 was excluded from the mortality burden analyses. From 2007 to 2023, an estimated 6,576 deaths (95% CI: 3,990, 9,155) annually across the contiguous U.S. were attributable to drought months and 10,465 deaths (95% CI: 6,642, 14,261) per year to wildfire smoke days, with an additional 469 deaths (95% CI: 256, 682) per year attributable to their concurrence. This corresponds to approximately 20.7 deaths (95%CI: 12.5, 28.8), 32.9 deaths (95%CI: 20.9, 44.8), 1.5 deaths (95%CI: 0.8, 2.2) per 1,000,000 individuals annually attributable to drought, wildfire smoke, and their concurrent added, respectively. Cause-specific mortality attributable to drought, wildfire smoke, and their concurrent added burden for the total population, as well as stratified by sex, age and SVI group, are shown in **Table S3**.

Attributable mortality rates for drought, wildfire smoke, and their concurrent day added burden varied across time and space, as shown in **Figure 4**, closely reflecting the spatiotemporal patterns of exposure in **Figure 1**. Mortality attributable to drought increased from 2007–2010 to 2011– 2015, then declined in 2016–2020, before rising again in 2021–2023. Drought-related mortality was initially concentrated in parts of the Pacific and southern U.S. It later remained elevated in California, Nevada, while high attributable mortality in the southern regions shifted westward into Texas and continued north into states like New Mexico, Oklahoma, Kansas, and Nebraska. Mortality then restricted to Washington and Oregon, and by 2021–2023, it affected broader areas across the Pacific and Mountain regions. For wildfire smoke, attributable mortality rates during 2021–2023 were substantially higher than in previous periods. The Northwest experienced increasing mortality in both intensity and geographic extent over time. In the East, attributable mortality rates were lowest during 2016–2020 but rose markedly in 2021–2023. Concurrent-day added mortality was also peaked in 2021–2023. In 2007–2010, elevated rates were concentrated in the South Atlantic and East South Central regions, then shifted northwestward into the West South Central region and showed increases in Wyoming, Wisconsin, and surrounding areas in 2011–2015. This burden declined markedly during 2016–2020 before rising again in 2021–2023, particularly across many areas in the West North Central and West South Central regions, as well as other scattered parts of the country.

**Figure 4.**
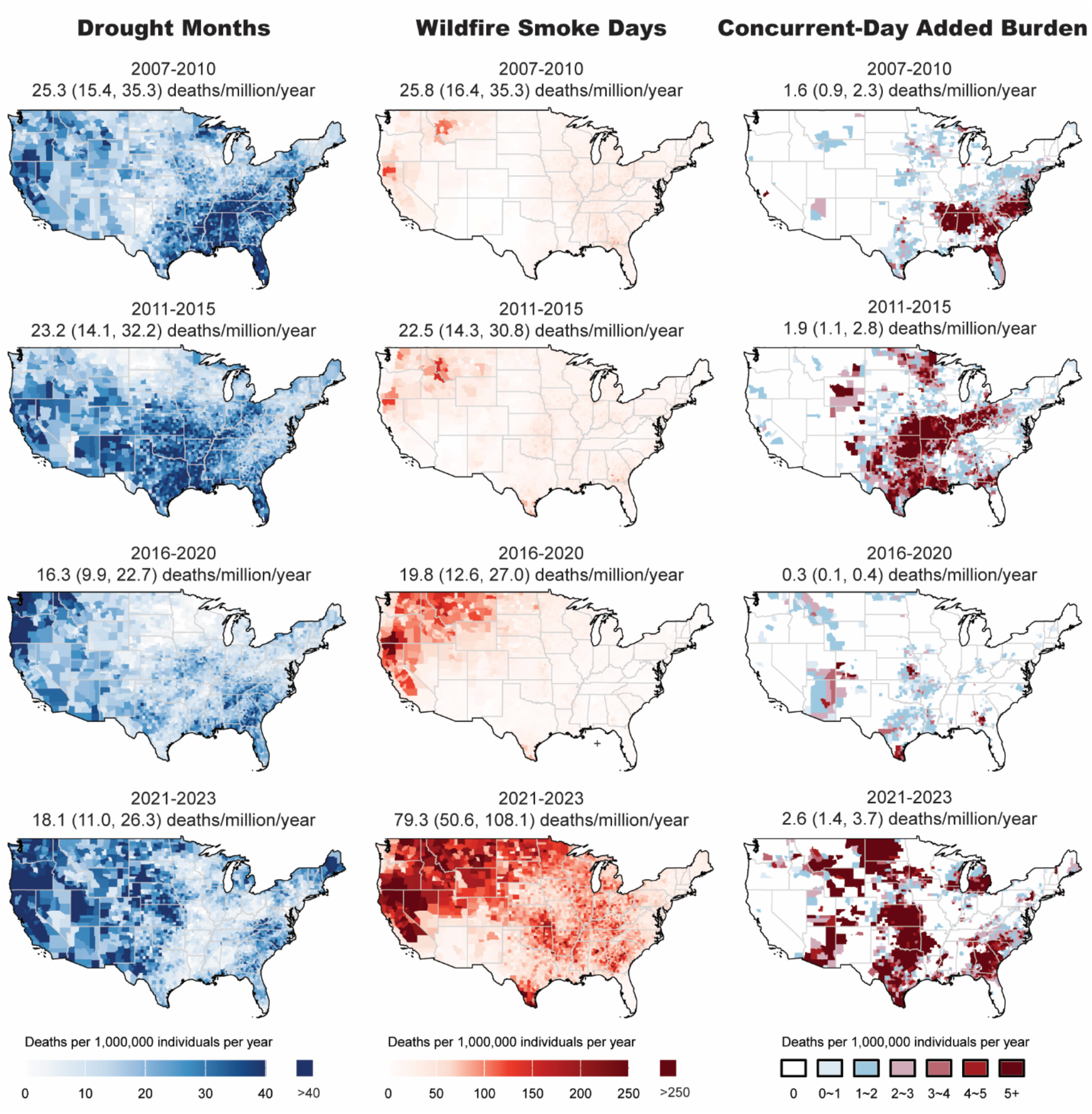
All-cause mortality rate attributable to drought months, wildfire smoke days and added effects of their concurrent days across 3,103 U.S. counties over four periods (2007–2010, 2011–2015, 2016–2020, and 2021–2023).

We found that mortality attributable to wildfire smoke [44.4 deaths (95%CI: 26.5, 62.1), 12.3 deaths (95%CI: 1.8, 22.7) per 1,000,000 individuals per year for high SVI and low SVI groups, respectively], and their concurrent added burden [2.6 deaths (95%CI: 1.6, 3.7), 0.7 deaths (95%CI: −0.1, 1.6) per 1,000,000 individuals per year for high SVI and low SVI groups, respectively] was consistently higher in high SVI counties compared to low SVI counties. This pattern may be attributable to both amplified health effects and higher baseline mortality rate in socially vulnerable populations. The pattern of concurrent added burden is also attributable to more exposure for socially vulnerable populations. The mortality attributable to drought was not significantly different [19.6 deaths (95%CI: 7.4, 31.7), 20.2 deaths (95%CI: 11.4, 29.1) per 1,000,000 individuals per year for high SVI and low SVI groups, respectively]. The county-level mortality attributable to drought, wildfire smoke and their concurrent-day added burden, stratified by SVI, were illustrated in **Figure 5**. We found some counties with high drought-attributable mortality and wildfire smoke-attributable mortality also experienced high concurrent-day added burden, especially for counties in high SVI group, indicating drought, wildfire smoke, and their added burden are concentrated in certain high SVI regions that need targeted interventions.

**Figure 5.**
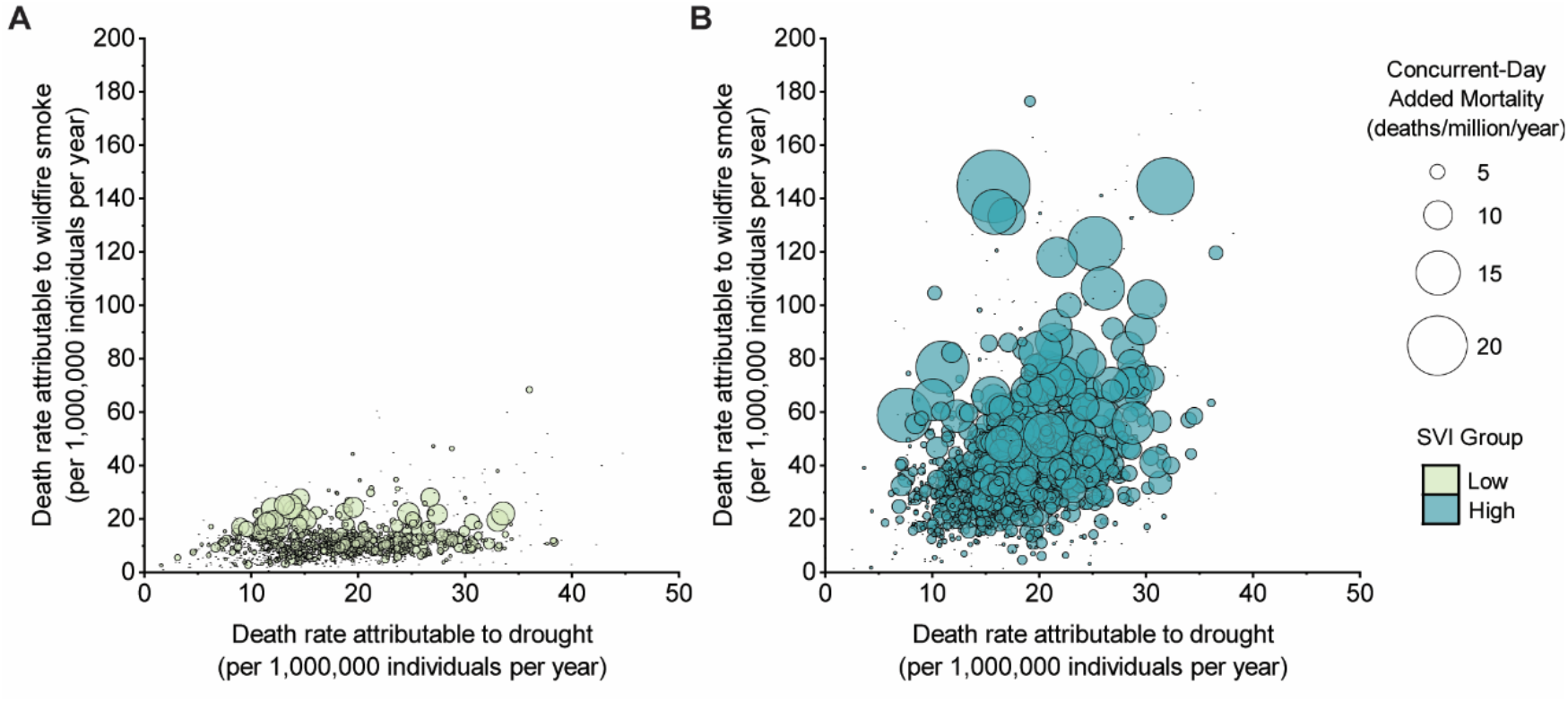
All-cause mortality rate attributable to drought months, wildfire smoke days and added effects of their concurrent days across 3,103 U.S. counties, 2007–2023, by Social Vulnerability Index (SVI) group. A. Low SVI counties (bottom 50%, n = 1551). B. High SVI counties (top 50%, n = 1552).

### Sensitivity analysis

We explored the association between cause-specific mortality and drought, wildfire smoke, and their concurrent added effect under alternative SPEI timescales (3, 6, and 12 months) and thresholds (−1.0, −1.1, −1.3, −1.4, and −1.5), as well as daily wildfire smoke PM_2.5_ thresholds (5 and 15 µg/m^3^), along with their attributable all-cause mortality under these metrics. The association was generally robust at the 1-month timescale of SPEI, but the effect of drought month and concurrent added effect attenuated and became non-significant at longer SPEI timescales (**Figures S3–6, Table S4**). The RRs of drought months, wildfire smoke days, and their concurrent-day added effect remained robust across a range of sensitivity analyses, including varying the degrees of freedom for key covariates, adding additional covariates, and testing alternative fixed effects (**Figures S7–9**). Comparing our TIAE model with the separate models that assessed drought and wildfire effects individually, as well as with the joint model that simultaneously evaluated drought, wildfire, and the concurrent added effect, we found that the individual effects of drought and wildfire were robust across models. However, in the joint model, the confidence interval for the concurrent added effect was wider than that observed in the TIAE model (**Figures S7–9**). We examined exposure periods from as short as the current month to as long as the current and previous 23 months, and found that the associations of the three exposures with all-cause mortality generally remained stable for longer exposure windows (**Figures S10–12**). Spatial and temporal randomization tests suggested that the model was not driven by spatial or temporal dependence due to misspecification (**Figure S13**).

## Discussion

This study characterizes the occurrence and co-occurrence of drought and wildfire smoke events across the contiguous U.S., evaluates their individual and concurrent added effect, and estimates the associated mortality burden. From 2006 to 2023, across 3,103 counties, we identified an annual average of 3,630 county-months of drought, 14,049 county-days of wildfire smoke, and 980 county-days of their concurrent. Both drought and wildfire smoke were significantly associated with increased all-cause mortality, and a significant added effect was identified when the two exposures occurred concurrently. We estimate that, on average, 6,576 (95% CI: 3,990, 9,155) annual deaths were attributable to drought months, 10,465 (95% CI: 6,642, 14,261) to wildfire smoke days, and an additional 469 (95% CI: 256, 682) to added effect of their concurrent days, with a higher burden observed in high-SVI counties. This study provides a novel national assessment of drought-attributable mortality burden and to assess the added effects and additional deaths associated with concurrent drought and wildfire smoke in the U.S., offering new insight into the epidemiological consequences of climate extremes and their concurrence. The TIAE model developed in this study enables assessments of the health impacts of concurrent climate-related events, enabling more integrated risk assessments and informing targeted public health interventions.

Previous research suggests that drought may affect health indirectly by disrupting environmental and social systems, such as nutrition, water quality, air pollution, and mental well-being ^32-35^. A possible explanation proposed in prior studies is that sustained drought can erode economic stability and living conditions, potentially leading to chronic psychological stress ^36^. This stress may, in turn, impair endocrine and immune function, increasing the risk of cardiovascular diseases ^37^. These evidences and pathways may help explain the associations observed in this study between drought exposure and elevated mortality from all causes, non-external, endocrine, nutritional and metabolic diseases, and CVDs. Consistent with our findings, some studies have linked severe drought to increased all-cause mortality, cardiovascular mortality, and hospital admissions in the U.S. ^12,13^, yet national estimates of drought-attributable deaths remain scarce, underscoring the contribution of this study in addressing this gap.

Wildfire smoke, particularly PM_2.5_, may impact health by inducing systemic inflammation, oxidative stress, and endothelial dysfunction ^38-41^. These processes can contribute to cardiovascular and metabolic disorders, impair liver and digestive function, and disrupt neuroendocrine regulation. Such mechanisms may underline the observed associations between wildfire smoke exposure and increased mortality across multiple causes, including non-external, CVD, respiratory diseases, endocrine, nutritional, and metabolic diseases, digestive diseases and mental and behavioral disorders. Previous studies have also found significant associations between wildfire exposure and increases in these cause-specific mortalities ^17,42^, and have estimated a wide range of annual premature deaths attributable to wildfire PM_2.5_ in the U.S. ^17,20-23,35^, with our estimate align well with those for long-term exposure (**Table S5**).

As climate change drives more frequent and severe extreme events, understanding their patterns and health impacts is critical for developing public health and adaptation strategies. By filling the gap of the national-level estimates of drought-attributable mortality across the contiguous U.S., this study contributes to a more comprehensive understanding of the relative health burdens posed by major climate-related hazards. Based on our estimates, drought was associated with 6,576 deaths (95% CI: 3,990, 9,155) and wildfire smoke with 10,465 deaths (95% CI: 6,643, 14,261) annually in the contiguous U.S. from 2007 to 2023. In comparison, existing evidence from the Global Burden of Disease Study 2021 (GBD 2021) ^43^ estimated 88,977 deaths (95% CI: 75,720, 97,544) from low temperature, 8,194 deaths (95% CI: 3,048, 16,407) from high temperature, 66,714 deaths (95% CI: 40,698, 95,806) from ambient PM_2.5_, and 15,258 deaths (95% CI: 3,352, 26,175) from ambient ozone annually in the U.S. from 2007 to 2021, and Chu et al. ^44^ estimated 1,178 deaths (95% CI: 356, 2,000) attributable to floods annually in the U.S. from 2001 to 2020. This rough comparison with previous studies suggests that, in the U.S., the mortality burden attributable to drought is higher than that attributable to floods, slightly lower than that attributable to high temperature, while the burden attributable to wildfire smoke is slightly higher than that attributable to high temperature but lower than that attributable to ambient ozone. All of these are substantially lower than the mortality attributable to ambient PM_2.5_ and low temperature. These findings can help inform the prioritization of targeted mitigation and adaptation policies to multiple extreme weather and climate events in the U.S.

While individual extreme events are becoming more frequent under climate change, their concurrence is also increasing^45^, highlighting the growing need to investigate whether these co-occurring events pose additional health burdens. Concurrent exposure to drought and wildfire smoke posed a significant added mortality risk, associated with all-cause, non-accidental, and multiple specific causes, including cardiovascular, respiratory, endocrine, nutritional and metabolic diseases, and mental and behavioral disorders. Although direct evidence is limited, this may be explained by the combined impact of drought-related chronic stress and wildfire smoke-induced inflammation^33,36^. Stress can impair immune and neuroendocrine function ^46^, while PM_2.5_ triggers oxidative stress and endothelial dysfunction ^47^. Similar interaction have been reported for other extreme events co-occurred with extreme temperature, such as heat-wildfire^17,26^, extreme temperature-PM_2.5_^27,28,48^, heat-ozone^29^, along with evidence that compound drought and heatwaves elevate mortality risk in patients with COPD ^25^, which may help explain the added effect observed in this study. The additional burden associated with concurrent drought and wildfire smoke identified in this study suggests that extreme events may not only interact with temperature to produce added risks, as shown in previous studies, but also pose additional risks through broader interactions. Recognizing and incorporating these interactions is essential to reduce underestimation of health burden, and to guide future climate-resilient public health strategies targeting concurrent extreme events, such as developing compound early warning systems ^26^.

Disproportionately higher impacts were observed among socially vulnerable populations, especially in relation to wildfire smoke days and the added burden from its concurrence with drought. Counties with higher SVI often have higher baseline mortality rates, partly due to limited access to healthcare^49^. Socially vulnerable communities often live in lower-quality housing with poor sealing^50^, which facilitates the infiltration of outdoor pollutants into indoor environments. Lower income levels in these areas can also limit their ability to evacuate or invest in wildfire mitigation measures^51,52^. Drought poses a greater mental health threat to vulnerable populations living in rural and remote communities.^53^ This underscores the need to prioritize these communities in climate-health planning and resource allocation.

This study has several limitations. First, we used wildfire smoke PM_2.5_ as a proxy to identify wildfire smoke days, while other wildfire-related pollutants such as ozone and volatile organic compounds were not included ^54,55^. These pollutants may differ in spatial distribution, health effects, and interactions with drought. Due to the current lack of high-resolution and national wide data on wildfire-related pollutants other than PM_2.5_, a more comprehensive understanding of wildfire smoke exposure and its health effects, including its interaction with drought, is currently constrained but represent a critical area for further research. Second, non-wildfire smoke PM_2.5_ was estimated by subtracting wildfire-attributed PM_2.5_ from a separate total PM_2.5_ dataset ^56,57^, which may introduce uncertainty. More integrated datasets that clearly distinguish ambient PM_2.5_ sources are needed in the future. Third, we did not examine other compound extreme events, such as drought-heat, drought-flood, or wildfire smoke-heat. Future research is warranted to elucidate the health impacts of these compound events.

This study demonstrates significant health risks and substantial mortality burdens associated with both drought and wildfire smoke exposure, with their concurrence imposing an additional burden in the U.S. Counties with high social vulnerability have higher health risks and experienced a larger mortality burden, compared with those with low social vulnerability. These findings emphasize the need to account for both individual and concurrent climate hazards in health impact assessments, and to inform the development of integrated, region-specific public health strategies in response to a changing climate.

## Materials and Methods

### Study population

We included 3,100 counties across the contiguous U.S., along with 3 combined counties, for a total of 3,103 counties in our analysis ^58^. The combined counties were constructed to account for boundary changes that occurred throughout the studied period, ensuring spatial and temporal consistency over the study period. Population data was stratified by sex (male and female) and by age group (0–24, 25–64 and ≥65 years). Annual county-level population data, disaggregated by sex and age group from 2006 to 2023, were obtained from the Surveillance, Epidemiology, and End Results (SEER) Program of the National Cancer Institute ^59^. The Social Vulnerability Index (SVI) for each county was derived from the 2010, 2016 and 2022 Centers for Disease Control and Prevention/ Agency for Toxic Substances and Disease Registry (CDC/ATSDR) SVI datasets, each based on 5-year American Community Survey (ACS) estimates. For each county, we calculated the average of overall social vulnerability across the three datasets, based on the composite of multiple vulnerability themes. Counties were then classified into high SVI (top 50%, 1552 counties) and low SVI (bottom 50%, 1551 counties) groups.

### Mortality Data

We obtained monthly cause-specific mortality data from the National Center for Health Statistics, covering 3,103 contiguous U.S. counties between 2007 and 2023. We examined deaths from all-cause (any ICD-10 code), non-external (any ICD-10 code excluding V00–Y99), cardiovascular diseases (CVD, I00–I99), respiratory diseases (J00–J99), endocrine, nutritional and metabolic diseases (E00–E89), digestive diseases (K00–K95), and mental and behavioral disorders (F01– F99). Mortality data were stratified by sex and age group, and mortality rates were age-standardized to the 2000 U.S. standard population. For analysis assessing association of drought month, wildfire smoke day, and their concurrent day with mortality, we restricted the dataset to 2007–2022 because daily total PM_2.5_ concentration data necessary for these analyses were available through 2022. For attributable mortality estimation, we utilized mortality data spanning 2007–2023 to assess the burden over a longer period.

Monthly age-standardized death counts were estimated by multiplying age-standardized rates by the corresponding population and rounding to the nearest whole. Counties were excluded from the health effect and attributable mortality analysis if, in all months without concurrent drought and wildfire smoke exposure during the current month and the preceding 11 months from 2007 to 2022 (i.e., the subset of data analyzed in Equation 1), monthly age-adjusted cause-specific deaths counts rounded to zero. A total of 37,958,499 all-cause deaths from 3,103 U.S. counties spanning 2007– 2023 were included in the analysis, including 19,078,866 males and 18,879,633 females. Age-stratified analyses focused on two groups (25–64 and ≥ 65 years), as individuals aged 0–24 years accounted for only 2% of total deaths and were excluded in age-stratified analysis due to limited cases. Total counts of cause-specific and subgroup deaths included in this study are summarized in **Table S6**, with most causes covering more than 95% of the counties.

### Extreme Event Exposure

Drought conditions were represented by the Standardized Precipitation Evapotranspiration Index (SPEI)^30^, which accounts for climatic water supply and demand simultaneously and thus provides a more robust and accurate measure than considering precipitation alone. Specifically, we first obtained monthly mean meteorological records at the resolution of 0.1° (∼9 km × 9 km) in the contiguous U.S. during 2006–2023 from ERA5-Land^60^. Next, we calculated the monthly potential evapotranspiration using historical records of maximum and minimum temperatures, dewpoint temperature, wind speed, solar radiation, air pressure, latitude, and elevation and the FAO-56 Penman-Monteith equation ^61^. Lastly, the monthly SPEI at the timescale of 1, 3, 6, and 12 months at the same spatial resolution was calculated using potential evapotranspiration and precipitation data. The detailed calculation method was described in our previous work ^62^. Gridded population data over the same time span at a 1 km × 1 km resolution for each year, sourced from the LandScan Global dataset ^63^ were used to generate population-weighted, county-level monthly averages of SPEIs. For our main analysis, we used the 1-month SPEI that captures short-term meteorological drought conditions. We identified county-months with a 1-month SPEI below −1.2 as drought months, approximately corresponding to the 10^th^ percentile during our studied period and indicating severe drought. The 10^th^ percentile has previously been used to identify drought based on SPEI and Standardized Precipitation Index (SPI) ^31,32^. Given that drought can be characterized over multiple timescales and that previous research has applied different thresholds such as −1.0 and −1.3 to identify drought ^14,62^, we also conducted analyses using 3, 6, and 12 months SPEI and with alternative thresholds at −1.0, −1.1, −1.3, −1.4, and −1.5 to assess the robustness of our findings. The average county-level SPEIs during the studied period were shown in **Figure S14**.

County-level daily population weighted wildfire smoke PM_2.5_ concentration data in the contiguous U.S. from 2006 to 2023 were obtained from a previous study^57^. These estimates were generated using a gradient boosted tree model trained on surface PM_2.5_ measurements from U.S. Environmental Protection Agency data and satellite inputs. The model is validated (R^2^ = 0.67) and specifically designed to isolate wildfire-related PM_2.5_ and provides consistent national coverage. We defined wildfire smoke day as a day the wildfire smoke PM_2.5_ concentration reached or exceeded 10 μg/m^3^, approximately corresponding to 99^th^ percentile of daily wildfire smoke PM_2.5_ concentration from 2006 to 2023. We also explore other thresholds including 5 and 15 μg/m^3^ (WHO 2021 air quality guideline for 24h PM_2.5_) to assess the robustness of our findings. The county-level wildfire PM_2.5_ concentration during the studied period were shown in **Figure S15**.

The concurrent drought and wildfire smoke days were defined as wildfire smoke days occurring within a drought month. We summarized the number of drought months, wildfire smoke days, and concurrent exposure days for each month. To capture long-term exposure, we calculated the counts of drought months, wildfire smoke days, and concurrent exposure days over the current and preceding 11 months for each county-month from 2007 to 2023. Data from 2006 was used for calculations but excluded from health effect analysis, as only December had a full 11 months of prior data to support long-term exposure calculation.

### Covariates

We considered several covariates to ensure that the effects of drought months, wildfire smoke days, and the added effect of their concurrence were not confounded by other factors. Covariates included in the main analyses comprised ambient temperature, precipitation ^64^, NDVI ^65^, and non-wildfire PM_2.5_ concentrations. Sensitivity analyses additionally considered relative humidity and snow water equivalence ^66^, and model comparison additionally considered SPEI and wildfire smoke PM_2.5_ concentrations (detailed in the sensitivity analysis section). These variables were selected based on their known or potential roles as confounders in the relationship between extreme events exposures (i.e., drought or wildfire smoke) and mortality. Each was represented as a 12-month moving average, calculated for each month from 2007 to 2022 using data from the current and preceding 11 months, to align with the long-term exposure window applied for extreme events. Statistical descriptions on these covariates were shown in **Table S7**.

Daily maximum and minimum air temperature, and water vapor pressure data at a 1 km × 1 km resolution from 2006 to 2022 were obtained from the Daymet Version 4 R1 dataset ^67^. Daily mean temperature was calculated as the average of daily maximum and minimum temperatures. Relative humidity was derived from daily water vapor pressure and saturated water vapor pressure, with the latter estimated from mean temperature based on the Clausius-Clapeyron relation, implemented via the empirical Tetens formula^68^. Then we used the a 1 km × 1 km population data for each year from the LandScan Global dataset ^63^ to generate population-weighted, county-level daily averages of ambient temperature and relative humidity. The daily values were aggregated into 12-month moving averages to estimate long-term exposure. Monthly average precipitation and snow water equivalent at a 1 km × 1 km resolution from 2006 to 2022 were obtained from the Daymet Version 4 R1 dataset^69^. Population-weighted, county-level monthly means were calculated and similarly aggregated using 12-month moving averages.

Monthly NDVI data at a 1 km × 1 km resolution from 2006 to 2022 were obtained from the MODIS dataset ^70^. Low-quality NDVI values were identified and removed based on associated quality control flags according to MODIS Vegetation Index User’s Guide^62^, including indicators of poor overall quality, cloud contamination, and precipitation. Missing values were then imputed using an Empirical Orthogonal Function (EOF) gap-filling method (R package *gapfill*) to ensure spatial and temporal continuity ^71^. After gap filling, population-weighted, county-level NDVI values were aggregated by month and then averaged over 12-month exposure windows.

Daily PM_2.5_ concentration data at a 1 km × 1 km resolution from 2006 to 2022 were obtained from the USHighAirPollutants (USHAP) dataset. This dataset were generated using a deep learning that integrated big data from satellites, models, and surface observations, with a cross-validated R^2^ at 0.80 ^56^. Population-weighted daily PM_2.5_ exposure was calculated at the county level using the same approach as for temperature data. To estimate non-wildfire PM_2.5_ concentrations, wildfire smoke-related PM_2.5_ was subtracted from total PM_2.5_ exposure for each county and day, with negative values set to zero. Daily exposures were then aggregated to monthly averages, which were used to calculate 12-month moving averages to represent long-term non-wildfire smoke PM_2.5_ exposure and wildfire smoke PM_2.5_ exposure. We also calculated the 12-month moving average county-level population-weighted SPEI using the 1-month SPEI mentioned in the previous section.

### Statistical Analysis

We developed a novel approach, termed the Two-Step Individual and Added Effect Estimation (TIAE) Model, to assess the individual effects of extreme events and to isolate any additional effect arising from their concurrence under long-term exposure. This method employs a two-step process of data filtering and analysis to split the individual effects and the added effect attributable to their concurrence, thereby addressing potential issues arising from correlations between extreme events and their concurrent exposures. The core principle is that, conditional on the individual effects of each exposure, any additional association between concurrent exposure and the health outcome indicates an added effect beyond individual contribution. Our model proceeds in two steps as illustrated in **Figure S1**. In the first step, we analyzed only observations without concurrent events to avoid confounding the estimation of individual effect, estimating the association between the number of exposure days (or months) for each extreme event and the health outcome using a generalized linear model. In the second step, we applied this model to estimate the expected counts of the health outcome for all observations, treating these estimations as an offset in a subsequent model that examined the association between concurrent days and the health outcome, thus isolating the added effect attributable to concurrent exposure.

All models were estimated using quasi-Poisson regression, which accounts for overdispersion in monthly mortality counts. Given that our dataset includes observations from over 3,000 counties, spanning multiple years and aggregated at a monthly resolution, we incorporated fixed-effects terms for counties, year-months, and state-years in the first-step model to control for unmeasured spatial, temporal and spatiotemporal confounding. Fixed-effect groups (counties, year-months, and state-years) with all-zero age-adjusted death counts in the first-step model were excluded to prevent estimation errors in the fixed-effects model. To account for potential correlation of observations within counties over time, we applied cluster-robust standard errors at the county level. The first-step model only includes county-months without concurrent days over the current and past 11 months, and is specific as follows:

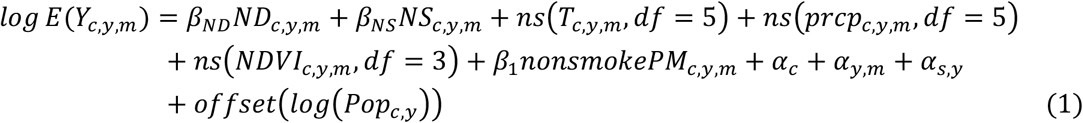

where *Y*_*c,y,m*_ is the number of deaths in county *c*, in month *m* of year *y*; *ND*_*c,y,m*_ is the counts of drought months of the current and previous 11 months in county *c*, in month *m* of year *y*; *β*_*ND*_ is the coefficient for drought exposure months; *NS*_*c,y,m*_ is the counts of wildfire smoke days of the current and previous 11 months in county *c*, in month *m* of year *y*; *β*_*NS*_ is the coefficient for wildfire smoke exposure days; *T*_*c,y,m*_ (unit: ℃) is the moving averages of ambient temperature of the current and previous 11 months in county *c*, in month *m* of year *y*, included as a covariate using a natural cubic spline with five degrees of freedom; *prcp*_*c,y,m*_ (unit: mm) is the moving averages of precipitation of the current and previous 11 months in county *c*, in month *m* of year *y*, included as a covariate using a natural cubic spline with five degrees of freedom; *NDVI*_*c,y,m*_ is the moving averages of NDVI of the current and previous 11 months in county *c*, in month *m* of year *y*, included as a covariate using a natural cubic spline with three degrees of freedom; *nonsmokePM*_*c,y,m*_ (unit: µg/m^3^) is the moving averages of non-wildfire smoke PM_2.5_ concentration of the current and previous 11 months in county *c*, in month *m* of year *y*; *β*_*1*_ is the coefficient for non-wildfire smoke PM_2.5_ exposure; *α*_*c*_ is the fix effect term for county *c*; *α*_*y,m*_ is the combined fix-effect term for year *y* and month of the year *m*; *α*_*s,y*_ is the combined fix-effect term for state *s* and year *y*; and *Pop*_*c,y*_ is the number of population in county *c* in year *y*, included as an offset to standardize for varying population sizes across counties and years.

The second step model applied to all county-months to estimate the added effect of concurrent days, was specified as follows:

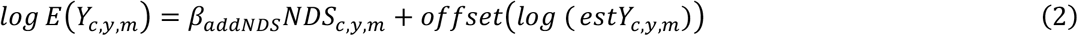

where *estY*_*c,y,m*_ is the number of estimated deaths without concurrent drought and wildfire smoke days in county *c*, in month *m* of year *y* estimated based on the first-step model; *NDS*_*c,y,m*_ is the counts of concurrent drought and wildfire smoke days of the current and previous 11 months in county *c*, in month *m* of year *y*; and *β*_*addNDS*_ is the coefficient for added effect of concurrent exposure days. The relative risk (RR) was further calculated using the following equation:

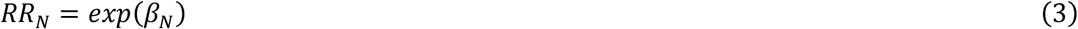

*RR*_*N*_ is the relative risk associated with per increase of drought month (*RR*_*ND*_) or wildfire smoke day (*RR*_*NS*_) in the current and previous 11 months, or the added effect per concurrent day increase in the current and previous 11 months (*RR*_*addNDS*_); and *β*_*N*_ is *β*_*ND*_, *β*_*NS*_,or *β*_*addNDS*_, respectively.

Using the TIAE model and long-term exposure and mortality data from 2007 to 2022, we estimated the RRs of cause-specific mortality associated with drought months, wildfire smoke days, and the added effect of their concurrence in the total population and stratified by age group, sex, and SVI groups. The difference between groups in RRs (*RR*_*diff*_) was assessed using the following equation:

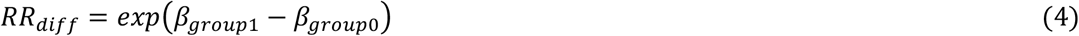

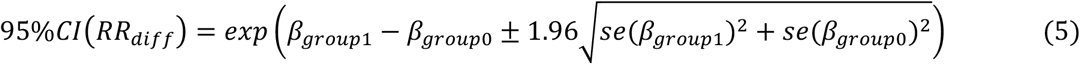

where *β*_*group1*_ is the coefficient for the comparison group, *β*_*group0*_ is the coefficient for the reference group, and *se(β*_*group1*_) and *se(β*_*group0*_) are the standard errors of *β*_*group1*_ and *β*_*group0*_. All analyses were conducted in R version 4.5.0 using the *fixest* package.

### Mortality Burden

The attributable fraction (AF), defined as the fraction of all cases of a health outcome that is attributable to a specific exposure ^72^, were estimated using the following equation:

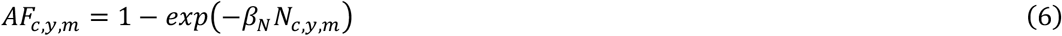

where *AF*_*c,y,m*_ is the AF due to drought months, wildfire smoke days, or their concurrent-day added effect in county *c*, in month *m* of year *y*; *N*_*c,y,m*_ is the count of drought months (*ND*_*c,y,m*_), wildfire smoke days (*NS*_*c,y,m*_), or their concurrent days (*NDS*_*c,y,m*_) of the current and previous 11 months in county *c*, in month *m* of year *y*. The attributable deaths and death rate were estimated using the following equations:

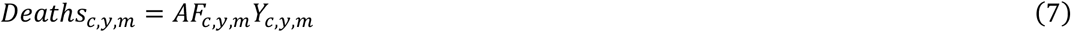

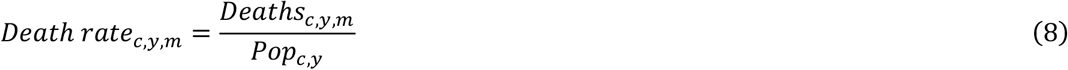

where *Deaths*_*c,y,m*_ is the deaths attributable to drought months, wildfire smoke days, or their concurrent-day added effect in county *c*, in month *m* of year *y*; *Death rate*_*c,y,m*_ is the death rate attributable to drought months, wildfire smoke days, or their concurrent-day added effect in county *c*, in month *m* of year *y*.

We estimated the total number of cause-specific deaths attributable to drought months, wildfire smoke days, and the additional burden due to their concurrence for the total population, as well as stratified by age, sex and SVI groups from 2007 to 2023. We further estimated county-level attributable deaths from all causes across four periods (2007–2010, 2011–2015, 2016–2020, and 2021–2023) using the RR for total population all-cause mortality to assess temporal trends in the attributable burden. Additionally, we examined differences across social vulnerability levels by applying SVI-specific RRs to each county to estimate attributable deaths by SVI group.

### Sensitivity Analysis

To understand how associations with cause-specific mortality and attributable deaths vary under different metrics, we evaluated alternative SPEI timescales (3, 6, and 12 months) and thresholds (−1.0, −1.1, −1.3, −1.4, and −1.5), along with daily wildfire smoke PM_2.5_ thresholds (5 and 15 µg/m^3^).

To test the robustness of our results, we performed several sensitivity analyses: (1) We adjusted the degrees of freedom for covariates, including temperature (df = 4 or 6), precipitation (df = 4 or 6), NDVI (df = 5), and non-wildfire smoke PM_2.5_ (df = 3). (2) We added additional covariates each at a time, including relative humidity and snow water equivalent, each modeled with natural cubic splines with three degrees of freedom. (3) We tested alternative fixed effects by including state-by-month combinations. Details of these sensitivity models are provided in **Table S8**.

To assess how different modeling strategies might influence estimates of both individual and added effects, we compared our TIAE model with two more traditional approaches. The first was a joint effect model, which simultaneously incorporated drought months, wildfire smoke days, and their concurrent into a generalized linear model, without the stepwise process. The second approach used separate models that evaluated individual effects of drought months and wildfire smoke days, without accounting for concurrent exposure. In these separate analyses, the drought month effect model additionally adjusted for the 12-month moving average wildfire smoke PM_2.5_, while the wildfire smoke day effect model adjusted for the 12-month moving average SPEI. Details of these comparison models are also presented in **Table S8**.

In addition, we expanded the exposure lag window from the current month and the preceding 11 months (lag 0–11) in our main analysis to a broader range of lag 0 to lag 0–23 months to test the lag pattern of both individual and added effect.

To exam the potential model misspecification, we also conducted spatial and temporal randomization tests as commonly used in previous studies ^17,58^. For the spatial test, we randomized county identifiers 2,000 times while preserving the year and month. For the temporal test, we randomized year and month 2,000 times while holding county identifiers constant. If the associations we originally observed become insignificant under these randomizations, it suggests that our findings are unlikely to be driven by spurious spatial or temporal dependencies.

## Data Availability

Monthly cause-specific mortality data can be requested from the National Center for Health Statistics (https://www.cdc.gov/nchs/index.htm). Exposure, population, and Social Vulnerability Index (SVI) data are publicly available or derived from publicly accessible sources with additional processing. Standardized precipitation evapotranspiration index (SPEI) data used in this study are available upon reasonable request from the corresponding author, Code used in this study will be available at https://github.com/CHENlab-Yale/Drought-Wildfire-AddedEffect after publication.

## Resource availability

### Materials availability

This study did not generate new unique materials/reagents.

## Funding and Acknowledgements

Y.H. was supported by the Li Foundation Climate Change Fellowship Program at the Yale Center on Climate Change and Health.

## Author Contributions

Conceptualization, Resources, and Supervision: K.C. Methodology, Formal analysis, Visualization, and Writing – Original Draft: Y.H. Data curation: Y.H., L.C., and P.W. Writing – Review & Editing: K.C., L.C., P.W., A.M.A., and M.Q.

## Supplemental Information

Supplemental information includes Figures S1–15, and Tables S1–8.

