## Supplemental Information for "Mortality Attributable to Drought, Wildfire Smoke, and Their Concurrent Added Effects in the Contiguous United States"

**Table S1 Number of drought months, wildfire smoke days, and their concurrent days under different event metrics, 2006–2023.**

| | Event Metrics | | | Number of Event Days or Months | | |
|---|---|---|---|---|---|---|
| | Daily Wildfire Smoke $PM_{2.5}$ | SPEI Timescale | Monthly SPEI | Drought Months | Wildfire Smoke Days | Co-occurrence Days |
| 1 | ≥ 10 μg/m$^3$ | SPEI01 | < -1.0 | 98414 | 252883 | 27512 |
| 2 | ≥ 10 μg/m$^3$ | SPEI01 | < -1.1 | 80932 | 252883 | 22311 |
| 3* | ≥ 10 μg/m$^3$ | SPEI01 | < -1.2 | 65332 | 252883 | 17632 |
| 4 | ≥ 10 μg/m$^3$ | SPEI01 | < -1.3 | 51496 | 252883 | 13661 |
| 5 | ≥ 10 μg/m$^3$ | SPEI01 | < -1.4 | 39382 | 252883 | 10495 |
| 6 | ≥ 10 μg/m$^3$ | SPEI01 | < -1.5 | 28919 | 252883 | 7744 |
| 7 | ≥ 10 μg/m$^3$ | SPEI03 | < -1.0 | 95143 | 252883 | 39158 |
| 8 | ≥ 10 μg/m$^3$ | SPEI03 | < -1.1 | 79126 | 252883 | 33006 |
| 9 | ≥ 10 μg/m$^3$ | SPEI03 | < -1.2 | 64953 | 252883 | 27535 |
| 10 | ≥ 10 μg/m$^3$ | SPEI03 | < -1.3 | 52162 | 252883 | 22467 |
| 11 | ≥ 10 μg/m$^3$ | SPEI03 | < -1.4 | 41133 | 252883 | 18020 |
| 12 | ≥ 10 μg/m$^3$ | SPEI03 | < -1.5 | 31634 | 252883 | 13848 |
| 13 | ≥ 10 μg/m$^3$ | SPEI06 | < -1.0 | 87329 | 252883 | 45494 |
| 14 | ≥ 10 μg/m$^3$ | SPEI06 | < -1.1 | 72677 | 252883 | 37462 |
| 15 | ≥ 10 μg/m$^3$ | SPEI06 | < -1.2 | 59550 | 252883 | 30371 |
| 16 | ≥ 10 μg/m$^3$ | SPEI06 | < -1.3 | 48223 | 252883 | 24326 |
| 17 | ≥ 10 μg/m$^3$ | SPEI06 | < -1.4 | 38067 | 252883 | 19030 |
| 18 | ≥ 10 μg/m$^3$ | SPEI06 | < -1.5 | 29445 | 252883 | 14505 |
| 19 | ≥ 10 μg/m$^3$ | SPEI012 | < -1.0 | 82319 | 252883 | 31255 |
| 20 | ≥ 10 μg/m$^3$ | SPEI012 | < -1.1 | 68769 | 252883 | 25249 |
| 21 | ≥ 10 μg/m$^3$ | SPEI012 | < -1.2 | 56452 | 252883 | 20054 |
| 22 | ≥ 10 μg/m$^3$ | SPEI012 | < -1.3 | 45086 | 252883 | 15569 |
| 23 | ≥ 10 μg/m$^3$ | SPEI012 | < -1.4 | 35232 | 252883 | 11577 |
| 24 | ≥ 10 μg/m$^3$ | SPEI012 | < -1.5 | 27017 | 252883 | 8185 |
| 25 | ≥ 5 μg/m$^3$ | SPEI01 | < -1.2 | 65332 | 795546 | 63957 |
| 26 | ≥ 5 μg/m$^3$ | SPEI03 | < -1.2 | 64953 | 795546 | 88368 |
| 27 | ≥ 5 μg/m$^3$ | SPEI06 | < -1.2 | 59550 | 795546 | 90011 |
| 28 | ≥ 5 μg/m$^3$ | SPEI012 | < -1.2 | 56452 | 795546 | 59243 |
| 29 | ≥ 15 μg/m$^3$ | SPEI01 | < -1.2 | 65332 | 104342 | 5200 |
| 30 | ≥ 15 μg/m$^3$ | SPEI03 | < -1.2 | 64953 | 104342 | 10656 |
| 31 | ≥ 15 μg/m$^3$ | SPEI06 | < -1.2 | 59550 | 104342 | 13321 |
| 32 | ≥ 15 μg/m$^3$ | SPEI012 | < -1.2 | 56452 | 104342 | 8768 |

* Main model.

**Table S2 Relative risk (RR and its 95% CI) and RR differences of monthly cause-specific mortality associated with per additional month of drought, per additional day of wildfire smoke, and per addition day of their concurrent during current and previous 11 months for individuals from different subgroups.**

| Cause | Group | Drought Months | | Wildfire Smoke Days | | Added Effect of Concurrent Days | |
|---|---|---|---|---|---|---|---|
| | | RR (95% CI) | Differences in RR (95% CI) | RR (95% CI) | Differences in RR (95% CI) | RR (95% CI) | Differences in RR (95% CI) |
| All-cause | Total | 1.0029 (1.0018, 1.0041)* | -- | 1.0013 (1.0008, 1.0018)* | -- | 1.0009 (1.0005, 1.0013)* | -- |
| | Males | 1.0032 (1.0018, 1.0045)* | Reference | 1.0014 (1.0009, 1.0020)* | Reference | 1.0008 (1.0002, 1.0013)* | Reference |
| | Females | 1.0027 (1.0014, 1.0039)* | 0.9995 (0.9977, 1.0013) | 1.0012 (1.0007, 1.0016)* | 0.9997 (0.9991, 1.0004) | 1.0010 (1.0004, 1.0016)* | 1.0002 (0.9994, 1.0010) |
| | 25–64 yrs | 1.0041 (1.0024, 1.0058)* | Reference | 1.0016 (1.0009, 1.0023)* | Reference | 1.0015 (1.0007, 1.0023)* | Reference |
| | ≥ 65 yrs | 1.0026 (1.0014, 1.0039)* | 0.9985 (0.9964, 1.0007) | 1.0010 (1.0006, 1.0015)* | 0.9995 (0.9986, 1.0003) | 1.0007 (1.0002, 1.0012)* | 0.9992 (0.9983, 1.0002) |
| | Low SVI | 1.0033 (1.0019, 1.0048)* | Reference | 1.0005 (1.0001, 1.0009)* | Reference | 1.0006 (0.9999, 1.0012) | Reference |
| | High SVI | 1.0023 (1.0009, 1.0038)* | 0.9990 (0.9970, 1.0011) | 1.0016 (1.0009, 1.0023)* | 1.0011 (1.0004, 1.0019)* | 1.0014 (1.0008, 1.0019)* | 1.0008 (0.9999, 1.0017) |
| Non-external | Total | 1.0029 (1.0016, 1.0041)* | -- | 1.0014 (1.0009, 1.0019)* | -- | 1.0012 (1.0008, 1.0016)* | -- |
| | Males | 1.0032 (1.0017, 1.0047)* | Reference | 1.0016 (1.0010, 1.0021)* | Reference | 1.0011 (1.0005, 1.0017)* | Reference |
| | Females | 1.0025 (1.0013, 1.0038)* | 0.9993 (0.9974, 1.0013) | 1.0012 (1.0007, 1.0017)* | 0.9996 (0.9989, 1.0004) | 1.0012 (1.0006, 1.0018)* | 1.0001 (0.9993, 1.0009) |
| | 25–64 yrs | 1.0041 (1.0023, 1.0060)* | Reference | 1.0021 (1.0013, 1.0030)* | Reference | 1.0025 (1.0016, 1.0033)* | Reference |
| | ≥ 65 yrs | 1.0026 (1.0013, 1.0040)* | 0.9985 (0.9962, 1.0008) | 1.0011 (1.0006, 1.0015)* | 0.9989 (0.9980, 0.9999)* | 1.0007 (1.0002, 1.0013)* | 0.9983 (0.9973, 0.9993)* |
| | Low SVI | 1.0033 (1.0018, 1.0048)* | Reference | 1.0006 (1.0001, 1.0010)* | Reference | 1.0004 (0.9998, 1.0011) | Reference |
| | High SVI | 1.0023 (1.0007, 1.0038)* | 0.9990 (0.9968, 1.0011) | 1.0017 (1.0010, 1.0024)* | 1.0011 (1.0003, 1.0020)* | 1.0019 (1.0013, 1.0025)* | 1.0015 (1.0006, 1.0024)* |
| Cardiovascular diseases | Total | 1.0020 (1.0006, 1.0034)* | -- | 1.0004 (1.0000, 1.0008)* | -- | 1.0013 (1.0006, 1.0020)* | -- |
| | Males | 1.0023 (1.0007, 1.0040)* | Reference | 1.0006 (1.0001, 1.0010)* | Reference | 1.0010 (1.0001, 1.0019)* | Reference |
| | Females | 1.0015 (0.9997, 1.0032) | 0.9991 (0.9967, 1.0015) | 1.0003 (0.9998, 1.0008) | 0.9998 (0.9991, 1.0004) | 1.0016 (1.0007, 1.0026)* | 1.0006 (0.9993, 1.0020) |
| | 25–64 yrs | 1.0013 (0.9990, 1.0037) | Reference | 1.0007 (1.0000, 1.0014) | Reference | 1.0044 (1.0029, 1.0059)* | Reference |
| | ≥ 65 yrs | 1.0020 (1.0005, 1.0035)* | 1.0007 (0.9979, 1.0035) | 1.0003 (0.9999, 1.0007) | 0.9996 (0.9987, 1.0004) | 1.0001 (0.9993, 1.0009) | 0.9958 (0.9941, 0.9975)* |
| | Low SVI | 1.0009 (0.9989, 1.0029) | Reference | 1.0001 (0.9996, 1.0006) | Reference | 1.0008 (0.9997, 1.0020) | Reference |
| | High SVI | 1.0022 (1.0005, 1.0040)* | 1.0013 (0.9987, 1.0040) | 1.0005 (1.0000, 1.0010) | 1.0004 (0.9996, 1.0012) | 1.0016 (1.0007, 1.0025)* | 1.0008 (0.9993, 1.0022) |
| Respiratory diseases | Total | 1.0018 (0.9992, 1.0043) | -- | 1.0006 (1.0001, 1.0012)* | -- | 1.0023 (1.0011, 1.0035)* | -- |
| | Males | 1.0019 (0.9988, 1.0049) | Reference | 1.0006 (0.9999, 1.0012) | Reference | 1.0023 (1.0007, 1.0040)* | Reference |
| | Females | 1.0012 (0.9983, 1.0042) | 0.9994 (0.9951, 1.0037) | 1.0006 (0.9999, 1.0012) | 1.0000 (0.9991, 1.0009) | 1.0021 (1.0003, 1.0038)* | 0.9997 (0.9973, 1.0021) |
| | 25–64 yrs | 1.0056 (1.0008, 1.0104)* | Reference | 1.0012 (1.0000, 1.0024) | Reference | 1.0055 (1.0023, 1.0087)* | Reference |
| | ≥ 65 yrs | 1.0011 (0.9986, 1.0037) | 0.9956 (0.9902, 1.0010) | 1.0004 (0.9999, 1.0009) | 0.9992 (0.9979, 1.0005) | 1.0011 (0.9997, 1.0025) | 0.9956 (0.9922, 0.9990)* |
| | Low SVI | 1.0060 (1.0018, 1.0101)* | Reference | 1.0000 (0.9992, 1.0008) | Reference | 1.0021 (1.0002, 1.0041)* | Reference |
| | High SVI | 0.9991 (0.9963, 1.0019) | 0.9931 (0.9882, 0.9981)* | 1.0009 (1.0002, 1.0016)* | 1.0009 (0.9998, 1.0020) | 1.0031 (1.0016, 1.0046)* | 1.0009 (0.9985, 1.0034) |

*$P$<0.05.

| Cause | Group | Drought Months | | Wildfire Smoke Days | | Added Effect of Concurrent Days | |
|---|---|---|---|---|---|---|---|
| | | RR (95% CI) | Differences in RR (95% CI) | RR (95% CI) | Differences in RR (95% CI) | RR (95% CI) | Differences in RR (95% CI) |
| Endocrine, nutritional and metabolic diseases | Total | 1.0047 (1.0014, 1.0080)* | -- | 1.0014 (1.0005, 1.0022)* | -- | 1.0033 (1.0016, 1.0050)* | -- |
| | Males | 1.0063 (1.0023, 1.0103)* | Reference | 1.0017 (1.0007, 1.0027)* | Reference | 1.0027 (1.0004, 1.0051)* | Reference |
| | Females | 1.0026 (0.9986, 1.0067) | 0.9963 (0.9907, 1.0020) | 1.0009 (0.9999, 1.0019) | 0.9992 (0.9978, 1.0005) | 1.0035 (1.0010, 1.0059)* | 1.0007 (0.9973, 1.0042) |
| | 25–64 yrs | 1.0061 (1.0014, 1.0108)* | Reference | 1.0009 (0.9999, 1.0019) | Reference | 1.0043 (1.0011, 1.0075)* | Reference |
| | ≥ 65 yrs | 1.0045 (1.0008, 1.0082)* | 0.9984 (0.9924, 1.0044) | 1.0013 (1.0004, 1.0022)* | 1.0004 (0.9990, 1.0017) | 1.0017 (0.9996, 1.0038) | 0.9974 (0.9936, 1.0012) |
| | Low SVI | 1.0029 (0.9977, 1.0082) | Reference | 1.0006 (0.9995, 1.0017) | Reference | 1.0014 (0.9985, 1.0043) | Reference |
| | High SVI | 1.0039 (1.0000, 1.0079) | 1.0010 (0.9944, 1.0076) | 1.0016 (1.0005, 1.0026)* | 1.0010 (0.9994, 1.0025) | 1.0051 (1.0030, 1.0072)* | 1.0036 (1.0001, 1.0072)* |
| Digestive diseases | Total | 1.0015 (0.9984, 1.0046) | -- | 1.0013 (1.0007, 1.0019)* | -- | 1.0019 (1.0000, 1.0039) | -- |
| | Males | 0.9987 (0.9943, 1.0032) | Reference | 1.0009 (1.0001, 1.0018)* | Reference | 1.0041 (1.0014, 1.0067)* | Reference |
| | Females | 1.0046 (1.0005, 1.0087)* | 1.0058 (0.9998, 1.0119) | 1.0017 (1.0008, 1.0026)* | 1.0007 (0.9995, 1.0020) | 0.9995 (0.9965, 1.0024) | 0.9954 (0.9915, 0.9994)* |
| | 25–64 yrs | 0.9998 (0.9950, 1.0045) | Reference | 1.0010 (1.0002, 1.0019)* | Reference | 1.0044 (1.0013, 1.0075)* | Reference |
| | ≥ 65 yrs | 1.0024 (0.9985, 1.0063) | 1.0026 (0.9965, 1.0088) | 1.0012 (1.0005, 1.0019)* | 1.0002 (0.9991, 1.0013) | 1.0008 (0.9982, 1.0034) | 0.9964 (0.9924, 1.0004) |
| | Low SVI | 1.0048 (0.9995, 1.0102) | Reference | 1.0008 (0.9998, 1.0018) | Reference | 0.9974 (0.9940, 1.0008) | Reference |
| | High SVI | 0.9996 (0.9959, 1.0033) | 0.9948 (0.9884, 1.0012) | 1.0014 (1.0008, 1.0021)* | 1.0006 (0.9994, 1.0018) | 1.0041 (1.0017, 1.0065)* | 1.0067 (1.0025, 1.0109)* |
| Mental and behavioral disorders | Total | 0.9998 (0.9949, 1.0048) | -- | 1.0025 (1.0013, 1.0038)* | -- | 1.0025 (1.0008, 1.0041)* | -- |
| | Males | 1.0012 (0.9953, 1.0071) | Reference | 1.0027 (1.0013, 1.0040)* | Reference | 0.9999 (0.9973, 1.0026) | Reference |
| | Females | 0.9993 (0.9939, 1.0047) | 0.9981 (0.9901, 1.0061) | 1.0026 (1.0013, 1.0039)* | 1.0000 (0.9981, 1.0019) | 1.0026 (1.0004, 1.0048)* | 1.0027 (0.9992, 1.0061) |
| | 25–64 yrs | 1.0053 (0.9911, 1.0198) | Reference | 1.0015 (0.9991, 1.0040) | Reference | 1.0030 (0.9976, 1.0085) | Reference |
| | ≥ 65 yrs | 1.0001 (0.9951, 1.0052) | 0.9949 (0.9799, 1.0100) | 1.0025 (1.0013, 1.0038)* | 1.0010 (0.9983, 1.0038) | 1.0014 (0.9996, 1.0033) | 0.9984 (0.9927, 1.0042) |
| | Low SVI | 1.0028 (0.9971, 1.0085) | Reference | 1.0014 (1.0001, 1.0026)* | Reference | 1.0027 (1.0001, 1.0052)* | Reference |
| | High SVI | 0.9985 (0.9919, 1.0051) | 0.9957 (0.9871, 1.0044) | 1.0028 (1.0010, 1.0045)* | 1.0014 (0.9993, 1.0035) | 1.0027 (1.0005, 1.0048)* | 1.0000 (0.9966, 1.0033) |

*$P<0.05$.

**Table S3 Annual cause-specific deaths attributable to drought months, wildfire smoke days, and added burden of concurrent days for individuals from different subgroups in the contiguous U.S., 2007-2023.**

| Cause | Group | Drought Months (95% CI) | Wildfire Smoke Days (95% CI) | Concurrent-Day Added Burden (95% CI) |
|---|---|---|---|---|
| All-cause | Total | 6576 (3990, 9155) | 10465 (6642, 14261) | 469 (256, 682) |
| | Males | 4123 (2377, 5864) | 6636 (4129, 9124) | 229 (67, 392) |
| | Females | 2599 (1401, 3795) | 4035 (2534, 5526) | 222 (99, 344) |
| | 25–64 yrs | 2140 (1242, 3034) | 3002 (1690, 4302) | 180 (86, 273) |
| | ≥ 65 yrs | 5049 (2625, 7466) | 7485 (4310, 10639) | 295 (86, 503) |
| | Low SVI | 2386 (1339, 3430) | 1450 (212, 2681) | 87 (-13, 186) |
| | High SVI | 3552 (1335, 5762) | 8058 (4808, 11278) | 479 (282, 676) |
| Non-external | Total | 5939 (3390, 8481) | 10249 (6501, 13970) | 556 (354, 757) |
| | Males | 3752 (2037, 5462) | 6568 (4176, 8940) | 281 (128, 434) |
| | Females | 2345 (1163, 3523) | 3915 (2395, 5426) | 248 (130, 366) |
| | 25–64 yrs | 1692 (926, 2455) | 3096 (1837, 4339) | 231 (151, 310) |
| | ≥ 65 yrs | 4934 (2498, 7363) | 7439 (4336, 10521) | 292 (86, 498) |
| | Low SVI | 2194 (1175, 3210) | 1614 (384, 2837) | 61 (-34, 156) |
| | High SVI | 3176 (982, 5361) | 7778 (4575, 10947) | 609 (422, 795) |
| Cardiovascular diseases | Total | 1366 (397, 2332) | 1024 (4, 2038) | 205 (100, 309) |
| | Males | 962 (289, 1632) | 792 (152, 1429) | 91 (5, 176) |
| | Females | 432 (-92, 953) | 315 (-179, 806) | 108 (44, 171) |
| | 25–64 yrs | 163 (-121, 444) | 299 (-6, 602) | 125 (82, 168) |
| | ≥ 65 yrs | 1304 (302, 2303) | 653 (-339, 1639) | 15 (-97, 127) |
| | Low SVI | 195 (-243, 630) | 95 (-392, 578) | 39 (-13, 90) |
| | High SVI | 1070 (242, 1895) | 778 (-39, 1589) | 173 (80, 266) |
| Respiratory diseases | Total | 372 (-165, 906) | 433 (47, 815) | 108 (53, 162) |
| | Males | 225 (-147, 595) | 227 (-42, 493) | 62 (17, 106) |
| | Females | 120 (-167, 405) | 178 (-33, 387) | 43 (7, 79) |
| | 25–64 yrs | 159 (23, 292) | 115 (-2, 229) | 36 (15, 56) |
| | ≥ 65 yrs | 235 (-298, 766) | 290 (-83, 660) | 47 (-13, 106) |
| | High SVI | -132 (-533, 265) | 378 (84, 670) | 97 (49, 143) |
| | Low SVI | 410 (126, 691) | -5 (-229, 216) | 32 (3, 60) |
| Endocrine, nutritional and metabolic diseases | Total | 499 (151, 845) | 550 (211, 885) | 81 (40, 122) |
| | Males | 389 (144, 632) | 402 (178, 624) | 38 (5, 71) |
| | Females | 122 (-68, 310) | 156 (-18, 328) | 38 (11, 65) |
| | 25–64 yrs | 177 (40, 312) | 99 (-9, 206) | 29 (7, 51) |
| | ≥ 65 yrs | 389 (67, 708) | 451 (126, 772) | 33 (-8, 73) |
| | Low SVI | 96 (-78, 268) | 94 (-70, 256) | 10 (-10, 30) |
| | High SVI | 291 (-1, 580) | 414 (143, 682) | 89 (52, 125) |
| Digestive diseases | Total | 122 (-132, 374) | 380 (203, 555) | 34 (-1, 69) |
| | Males | -61 (-273, 150) | 162 (10, 313) | 42 (15, 69) |
| | Females | 158 (16, 299) | 215 (100, 328) | -4 (-27, 19) |
| | 25–64 yrs | -8 (-163, 145) | 123 (19, 226) | 31 (10, 53) |
| | ≥ 65 yrs | 129 (-84, 340) | 254 (109, 397) | 9 (-21, 39) |
| | Low SVI | 120 (-13, 251) | 87 (-20, 193) | -13 (-31, 4) |
| | High SVI | -24 (-233, 182) | 270 (145, 394) | 52 (22, 82) |
| Mental and behavioral disorders | Total | -21 (-579, 531) | 973 (506, 1432) | 63 (21, 106) |
| | Males | 61 (-240, 358) | 468 (233, 700) | -1 (-32, 30) |
| | Females | -42 (-363, 275) | 518 (265, 767) | 35 (6, 63) |
| | 25–64 yrs | 57 (-98, 207) | 68 (-42, 174) | 7 (-6, 20) |
| | ≥ 65 yrs | 15 (-565, 589) | 1013 (526, 1493) | 35 (-11, 81) |
| | Low SVI | 117 (-124, 354) | 233 (25, 437) | 25 (1, 48) |
| | High SVI | -108 (-575, 352) | 584 (219, 941) | 44 (8, 79) |

**Table S4 Annual all-cause deaths attributable to drought months, wildfire smoke days, and added burden of concurrent days under different event metrics in the contiguous U.S., 2007-2023.**

| | Event Metrics | | | Annual Deaths (95% CI) | | |
|---|---|---|---|---|---|---|
| | Wildfire Smoke Day | SPEI Timescale | SPEI | Drought | Wildfire Smoke | Added Burden |
| 1 | ≥ 10 μg/m³ | SPEI01 | SPEI < -1.0 | 5445 (1278, 9598) | 9572 (4901, 14204) | 1821 (1562, 2079) |
| 2 | ≥ 10 μg/m³ | SPEI01 | SPEI < -1.1 | 6405 (3406, 9396) | 10287 (6420, 14128) | 1025 (786, 1264) |
| 3* | ≥ 10 μg/m³ | SPEI01 | SPEI < -1.2 | 6576 (3990, 9155) | 10465 (6643, 14261) | 469 (256, 682) |
| 4 | ≥ 10 μg/m³ | SPEI01 | SPEI < -1.3 | 5896 (3564, 8222) | 10479 (6683, 14249) | 202 (12, 392) |
| 5 | ≥ 10 μg/m³ | SPEI01 | SPEI < -1.4 | 4106 (2200, 6007) | 10182 (6421, 13918) | 305 (137, 473) |
| 6 | ≥ 10 μg/m³ | SPEI01 | SPEI < -1.5 | 2135 (493, 3772) | 9923 (6187, 13634) | 274 (130, 418) |
| 7 | ≥ 10 μg/m³ | SPEI03 | SPEI < -1.0 | 4039 (1170, 6900) | 10696 (5577, 15768) | 1222 (942, 1502) |
| 8 | ≥ 10 μg/m³ | SPEI03 | SPEI < -1.1 | 3379 (957, 5795) | 9450 (4838, 14024) | 1264 (1011, 1517) |
| 9 | ≥ 10 μg/m³ | SPEI03 | SPEI < -1.2 | 4177 (1994, 6353) | 9665 (5251, 14044) | 690 (474, 906) |
| 10 | ≥ 10 μg/m³ | SPEI03 | SPEI < -1.3 | 2366 (-237, 4958) | 9463 (5236, 13659) | 796 (600, 992) |
| 11 | ≥ 10 μg/m³ | SPEI03 | SPEI < -1.4 | 2359 (629, 4083) | 9422 (5308, 13506) | 548 (372, 724) |
| 12 | ≥ 10 μg/m³ | SPEI03 | SPEI < -1.5 | 1013 (-311, 2333) | 9307 (5258, 13327) | 677 (520, 834) |
| 13 | ≥ 10 μg/m³ | SPEI06 | SPEI < -1.0 | 1780 (-638, 4189) | 9529 (5084, 13939) | 791 (573, 1009) |
| 14 | ≥ 10 μg/m³ | SPEI06 | SPEI < -1.1 | 2141 (-307, 4579) | 10599 (5729, 15427) | 286 (90, 481) |
| 15 | ≥ 10 μg/m³ | SPEI06 | SPEI < -1.2 | 1850 (-176, 3867) | 10229 (5510, 14909) | 426 (249, 603) |
| 16 | ≥ 10 μg/m³ | SPEI06 | SPEI < -1.3 | 863 (-1291, 3006) | 10049 (5626, 14437) | 450 (293, 606) |
| 17 | ≥ 10 μg/m³ | SPEI06 | SPEI < -1.4 | 533 (-1283, 2340) | 9474 (5391, 13528) | 556 (418, 694) |
| 18 | ≥ 10 μg/m³ | SPEI06 | SPEI < -1.5 | 92 (-1432, 1608) | 9099 (5101, 13069) | 585 (465, 705) |
| 19 | ≥ 10 μg/m³ | SPEI012 | SPEI < -1.0 | 520 (-1289, 2320) | 4219 (2481, 5947) | 1060 (899, 1221) |
| 20 | ≥ 10 μg/m³ | SPEI012 | SPEI < -1.1 | -34 (-1663, 1587) | 9449 (5154, 13711) | 760 (623, 897) |
| 21 | ≥ 10 μg/m³ | SPEI012 | SPEI < -1.2 | -142 (-1702, 1409) | 9687 (5607, 13738) | 511 (389, 632) |
| 22 | ≥ 10 μg/m³ | SPEI012 | SPEI < -1.3 | -100 (-1449, 1241) | 9768 (5789, 13719) | 17 (-92, 125) |
| 23 | ≥ 10 μg/m³ | SPEI012 | SPEI < -1.4 | 303 (-1493, 2081) | 9767 (5921, 13587) | -23 (-120, 73) |
| 24 | ≥ 10 μg/m³ | SPEI012 | SPEI < -1.5 | 235 (-1397, 1849) | 9833 (6101, 13540) | -107 (-195, -19) |
| 25 | ≥ 5 μg/m³ | SPEI01 | SPEI < -1.2 | 5744 (3017, 8463) | 29099 (18363, 39701) | 658 (363, 953) |
| 26 | ≥ 5 μg/m³ | SPEI03 | SPEI < -1.2 | 3845 (1353, 6329) | 28712 (17120, 40148) | 1201 (909, 1493) |
| 27 | ≥ 5 μg/m³ | SPEI06 | SPEI < -1.2 | 2780 (399, 5149) | 23588 (15483, 31617) | 313 (69, 556) |
| 28 | ≥ 5 μg/m³ | SPEI012 | SPEI < -1.2 | -423 (-2303, 1444) | 26785 (15684, 37743) | 654 (476, 832) |
| 29 | ≥ 15 μg/m³ | SPEI01 | SPEI < -1.2 | 6248 (3835, 8655) | 5040 (3458, 6614) | 557 (429, 685) |
| 30 | ≥ 15 μg/m³ | SPEI03 | SPEI < -1.2 | 3298 (1334, 5257) | 4618 (2906, 6320) | 783 (635, 930) |
| 31 | ≥ 15 μg/m³ | SPEI06 | SPEI < -1.2 | 1696 (-200, 3585) | 4577 (2569, 6572) | 171 (55, 287) |
| 32 | ≥ 15 μg/m³ | SPEI012 | SPEI < -1.2 | -218 (-1703, 1259) | 4552 (2813, 6281) | 392 (316, 468) |

* Main model.

**Table S5 Estimated annual number of deaths attributable to long-term wildfire smoke $PM_{2.5}$ exposure in the contiguous U.S. in this and previous studies.**

| Cause | Year | Estimated Annual Attributable Deaths | Reference |
|---|---|---|---|
| All-cause | 2007-2023 | 10,465 (95% CI: 6,655, 14,274) | This study |
| Non-accidental | 2007-2020 | 11,415 (95% CI: 6,754, 16,075) | Ma et al. [1] |
| All-cause | 2006–2018 | 6,300 (95% CI: 4,800, 7,800) | O'Dell et al. [2] |
| All-cause | 2012–2014 | 4,000 (95% CI: 2,700, 5,300) | Pan et al. [3] |
| All-cause | 2000 | 7,000 to 28,000 | Ford et al. [4] |
| All-cause | 2008–2012 | 8,700 (95% CI: 5,800, 11,000) to 32,000 (95% CI: 9,800, 29,000) | Fann et al. [5] |
| All-cause | 2011-2020 | 15,800 (95% CI: 6900, 25300) | Qiu et al. [6] |

**Table S6 Summary of counties analyzed and number of deaths by causes and subgroups, 2006-2023.**

| Cause | ICD-10 Code | Subgroup | Number of Counties (%) | Number of Deaths |
|---|---|---|---|---|
| All causes | Any | Total | 3103 (100%) | 37,958,499 |
| | | Males | 3103 (100%) | 19,078,866 |
| | | Females | 3103 (100%) | 18,879,633 |
| | | 25–64 yrs | 3102 (99.97%) | 8,873,021 |
| | | ≥ 65 yrs | 3103 (100%) | 28,350,860 |
| Non-external | Any excluding V00-Y99 | Total | 3103 (100%) | 35,020,434 |
| | | Males | 3103 (100%) | 17,078,056 |
| | | Females | 3102 (99.97%) | 17,942,378 |
| | | 25–64 yrs | 3102 (99.97%) | 7,099,266 |
| | | ≥ 65 yrs | 3103 (100%) | 27,553,170 |
| Cardiovascular diseases | I00-I99 | Total | 3101 (99.94%) | 11,704,828 |
| | | Males | 3100 (99.9%) | 5,852,858 |
| | | Females | 3099 (99.9%) | 5,851,960 |
| | | 25–64 yrs | 3092 (99.6%) | 2,082,432 |
| | | ≥ 65 yrs | 3102 (99.97%) | 9,599,203 |
| Respiratory diseases | J00-J99 | Total | 3092 (99.6%) | 3,503,795 |
| | | Males | 3082 (99.3%) | 1,679,687 |
| | | Females | 3072 (99.0%) | 1,824,020 |
| | | 25–64 yrs | 3005 (96.8%) | 496,823 |
| | | ≥ 65 yrs | 3094 (99.7%) | 2,988,963 |
| Endocrine, nutritional and metabolic diseases | E00-E89 | Total | 3086 (99.5%) | 1,807,590 |
| | | Males | 3072 (99.0%) | 933,293 |
| | | Females | 3050 (98.3%) | 874,131 |
| | | 25–64 yrs | 3033 (97.7%) | 489,678 |
| | | ≥ 65 yrs | 3085 (99.4%) | 1,305,750 |
| Digestive diseases | K00-K95 | Total | 3056 (98.5%) | 1,327,386 |
| | | Males | 3039 (97.9%) | 699,202 |
| | | Females | 3001 (96.7%) | 627,993 |
| | | 25–64 yrs | 2993 (96.5%) | 517,431 |
| | | ≥ 65 yrs | 3061 (98.6%) | 803,753 |
| Mental and behavioral disorders | F01-F99 | Total | 3061 (98.6%) | 1,894,610 |
| | | Males | 3044 (98.1%) | 682,643 |
| | | Females | 2996 (96.6%) | 1,211,577 |
| | | 25–64 yrs | 2858 (92.1%) | 175,936 |
| | | ≥ 65 yrs | 3060 (98.6%) | 1,715,792 |

**Table S7 Statistical description on 12-month moving average of environmental factors across 3,103 U.S. counties, 2007-2022.**

| Environmental Factors | Mean | SD | Min | Median | IQR | Max |
|---|---|---|---|---|---|---|
| Ambient temperature (°C) | 13.1 | 4.5 | 0.5 | 12.9 | 6.8 | 26.9 |
| Relative humidity (%) | 63 | 9 | 21 | 67 | 7 | 85 |
| Precipitation (mm) | 89 | 37 | 0 | 92 | 52 | 297 |
| Normalized difference vegetation index (NDVI) | 0.50 | 0.12 | 0.09 | 0.52 | 0.18 | 0.76 |
| Wildfire $PM_{2.5}$ smoke (μg/m$^3$) | 0.50 | 0.61 | 0.00 | 0.33 | 0.42 | 24.69 |
| Non-wildfire $PM_{2.5}$ smoke (μg/m$^3$) | 7.9 | 2.0 | 2.0 | 7.8 | 2.6 | 22.1 |
| Snow water equivalent (kg/m$^2$) | 4.6 | 9.8 | 0.0 | 0.8 | 4.7 | 252.1 |
| Standardized Precipitation Evapotranspiration Index (SPEI) | 0.09 | 0.34 | -1.57 | 0.10 | 0.45 | 1.24 |

**Table S8 List of the sensitivity analysis and comparison models.**

| | Model Equations | Model Description |
|---|---|---|
| **Main** | $log\,E(Y_{c,y,m}) = \beta_{ND}ND_{c,y,m} + \beta_{NS}NS_{c,y,m} + ns(T_{c,y,m}, df = 5) + ns(prcp_{c,y,m}, df = 5) + ns(NDVI_{c,y,m}, df = 3) + \beta_1 nonsmokePM_{c,y,m} + \alpha_c + \alpha_{y,m} + \alpha_{s,y} + offset(log(Pop_{c,y}))$<br>$log\,E(Y_{c,y,m}) = \beta_{addNDS}NDS_{c,y,m} + offset(log\,(estY_{c,y,m}))$ | Main model |
| **SA_1** | $log\,E(Y_{c,y,m}) = \beta_{ND}ND_{c,y,m} + \beta_{NS}NS_{c,y,m} + ns(T_{c,y,m}, df = 4) + ns(prcp_{c,y,m}, df = 5) + ns(NDVI_{c,y,m}, df = 3) + \beta_1 nonsmokePM_{c,y,m} + \alpha_c + \alpha_{y,m} + \alpha_{s,y} + offset(log(Pop_{c,y}))$<br>$log\,E(Y_{c,y,m}) = \beta_{addNDS}NDS_{c,y,m} + offset(log\,(estY_{c,y,m}))$ | Adjusted degree of freedom of temperature (df = 4) |
| **SA_2** | $log\,E(Y_{c,y,m}) = \beta_{ND}ND_{c,y,m} + \beta_{NS}NS_{c,y,m} + ns(T_{c,y,m}, df = 6) + ns(prcp_{c,y,m}, df = 5) + ns(NDVI_{c,y,m}, df = 3) + \beta_1 nonsmokePM_{c,y,m} + \alpha_c + \alpha_{y,m} + \alpha_{s,y} + offset(log(Pop_{c,y}))$<br>$log\,E(Y_{c,y,m}) = \beta_{addNDS}NDS_{c,y,m} + offset(log\,(estY_{c,y,m}))$ | Adjusted degree of freedom of temperature (df = 6) |
| **SA_3** | $log\,E(Y_{c,y,m}) = \beta_{ND}ND_{c,y,m} + \beta_{NS}NS_{c,y,m} + ns(T_{c,y,m}, df = 5) + ns(prcp_{c,y,m}, df = 4) + ns(NDVI_{c,y,m}, df = 3) + \beta_1 nonsmokePM_{c,y,m} + \alpha_c + \alpha_{y,m} + \alpha_{s,y} + offset(log(Pop_{c,y}))$<br>$log\,E(Y_{c,y,m}) = \beta_{addNDS}NDS_{c,y,m} + offset(log\,(estY_{c,y,m}))$ | Adjusted degree of freedom of precipitation (df = 4) |
| **SA_4** | $log\,E(Y_{c,y,m}) = \beta_{ND}ND_{c,y,m} + \beta_{NS}NS_{c,y,m} + ns(T_{c,y,m}, df = 5) + ns(prcp_{c,y,m}, df = 6) + ns(NDVI_{c,y,m}, df = 3) + \beta_1 nonsmokePM_{c,y,m} + \alpha_c + \alpha_{y,m} + \alpha_{s,y} + offset(log(Pop_{c,y}))$<br>$log\,E(Y_{c,y,m}) = \beta_{addNDS}NDS_{c,y,m} + offset(log\,(estY_{c,y,m}))$ | Adjusted degree of freedom of precipitation (df = 6) |
| **SA_5** | $log\,E(Y_{c,y,m}) = \beta_{ND}ND_{c,y,m} + \beta_{NS}NS_{c,y,m} + ns(T_{c,y,m}, df = 5) + ns(prcp_{c,y,m}, df = 5) + ns(NDVI_{c,y,m}, df = 5) + \beta_1 nonsmokePM_{c,y,m} + \alpha_c + \alpha_{y,m} + \alpha_{s,y} + offset(log(Pop_{c,y}))$<br>$log\,E(Y_{c,y,m}) = \beta_{addNDS}NDS_{c,y,m} + offset(log\,(estY_{c,y,m}))$ | Adjusted degree of freedom of NDVI (df = 5) |
| **SA_6** | $log\,E(Y_{c,y,m}) = \beta_{ND}ND_{c,y,m} + \beta_{NS}NS_{c,y,m} + ns(T_{c,y,m}, df = 5) + ns(prcp_{c,y,m}, df = 5) + ns(NDVI_{c,y,m}, df = 3) + ns(nonsmokePM_{c,y,m}, df = 3) + \alpha_c + \alpha_{y,m} + \alpha_{s,y} + offset(log(Pop_{c,y}))$<br>$log\,E(Y_{c,y,m}) = \beta_{addNDS}NDS_{c,y,m} + offset(log\,(estY_{c,y,m}))$ | Adjusted non-wildfire smoke $PM_{2.5}$ for as a non-linear covariate (nature cubic spline with degree of freedom at 3) |
| **SA_7** | $log\,E(Y_{c,y,m}) = \beta_{ND}ND_{c,y,m} + \beta_{NS}NS_{c,y,m} + ns(T_{c,y,m}, df = 5) + ns(prcp_{c,y,m}, df = 5) + ns(NDVI_{c,y,m}, df = 3) + ns(swe_{c,y,m}, df = 3) + \beta_1 nonsmokePM_{c,y,m} + \alpha_c + \alpha_{y,m} + \alpha_{s,y} + offset(log(Pop_{c,y}))$<br>$log\,E(Y_{c,y,m}) = \beta_{addNDS}NDS_{c,y,m} + offset(log\,(estY_{c,y,m}))$ | Adjusted for the snow water equivalent |
| **SA_8** | $log\,E(Y_{c,y,m}) = \beta_{ND}ND_{c,y,m} + \beta_{NS}NS_{c,y,m} + ns(T_{c,y,m}, df = 5) + ns(prcp_{c,y,m}, df = 5) + ns(NDVI_{c,y,m}, df = 3) + ns(RH_{c,y,m}, df = 3) + \beta_1 nonsmokePM_{c,y,m} + \alpha_c + \alpha_{y,m} + \alpha_{s,y} + offset(log(Pop_{c,y}))$<br>$log\,E(Y_{c,y,m}) = \beta_{addNDS}NDS_{c,y,m} + offset(log\,(estY_{c,y,m}))$ | Adjusted for the relative humidity |

| | | |
|---|---|---|
| **SA_9** | $\log E(Y_{c,y,m}) = \beta_{ND}ND_{c,y,m} + \beta_{NS}NS_{c,y,m} + ns(T_{c,y,m}, df = 5) + ns(prcp_{c,y,m}, df = 5) + ns(NDVI_{c,y,m}, df = 3) + \beta_1 nonsmokePM_{c,y,m} + \alpha_c + \alpha_{y,m} + \alpha_{s,y} + \alpha_{s,m} + offset(\log(Pop_{c,y}))$<br>$\log E(Y_{c,y,m}) = \beta_{addNDS}NDS_{c,y,m} + offset(\log(estY_{c,y,m}))$ | Adjusted for alternative fixed effect of state-month |
| **Joint** | $\log E(Y_{c,y,m}) = \beta_{ND}ND_{c,y,m} + \beta_{NS}NS_{c,y,m} + \beta_{addNDS}NDS_{c,y,m} + ns(T_{c,y,m}, df = 5) + ns(prcp_{c,y,m}, df = 5) + ns(NDVI_{c,y,m}, df = 3) + \beta_1 nonsmokePM_{c,y,m} + \alpha_c + \alpha_{y,m} + \alpha_{s,y} + offset(\log(Pop_{c,y}))$ | Joint model with drought months, wildfire smoke days and their concurrent days in a single model |
| **Separate** | $\log E(Y_{c,y,m}) = \beta_{ND}ND_{c,y,m} + ns(T_{c,y,m}, df = 5) + ns(prcp_{c,y,m}, df = 5) + ns(NDVI_{c,y,m}, df = 3) + \beta_1 nonsmokePM_{c,y,m} + \beta_2 smokePM_{c,y,m} + \alpha_c + \alpha_{y,m} + \alpha_{s,y} + offset(\log(Pop_{c,y}))$<br>$\log E(Y_{c,y,m}) = \beta_{NS}NS_{c,y,m} + ns(T_{c,y,m}, df = 5) + ns(prcp_{c,y,m}, df = 5) + ns(NDVI_{c,y,m}, df = 3) + \beta_1 nonsmokePM_{c,y,m} + \beta_3 SPEI_{c,y,m} + \alpha_c + \alpha_{y,m} + \alpha_{s,y} + offset(\log(Pop_{c,y}))$ | Separate models to analyze health effects of drought month and wildfire smoke day exposure, respectively.<br>Drought model: adjusted for wildfire smoke $PM_{2.5}$<br>Wildfire smoke model: adjusted for SPEI. |

* Differences with the main model were marked red.

$Y_{c,y,m}$, the number of deaths in county *c*, in month *m* of year *y*;
$estY_{c,y,m}$, the number of estimated deaths without concurrent drought and wildfire smoke days in county *c*, in month *m* of year *y* for all county-months estimated by the first-step model;
$ND_{c,y,m}$, the counts of drought months of the current and previous 11 months in county *c*, in month *m* of year *y*;
$NS_{c,y,m}$, the counts of wildfire smoke days of the current and previous 11 months in county *c*, in month *m* of year *y*;
$NDS_{c,y,m}$, the counts of concurrent drought and wildfire smoke days of the current and previous 11 months in county *c*, in month *m* of year *y*;
$\beta_{ND}$, the coefficient for drought exposure months;
$\beta_{NS}$, the coefficient for wildfire smoke exposure days;
$\beta_{addNDS}$, the coefficient for added effect of concurrent exposure days;
$T_{c,y,m}$ (unit: °C), the moving averages of ambient temperature of the current and previous 11 months in county *c*, in month *m* of year *y*;
$prcp_{c,y,m}$ (unit: mm), the moving averages of precipitation of the current and previous 11 months in county *c*, in month *m* of year *y*;
$NDVI_{c,y,m}$, the moving averages of NDVI of the current and previous 11 months in county *c*, in month *m* of year *y*;
$swe_{c,y,m}$ (unit: $kg/m^2$), the moving averages of snow water equivalent of the current and previous 11 months in county *c*, in month *m* of year *y*;
$RH_{c,y,m}$ (unit: %), the moving averages of relative humidity of the current and previous 11 months in county *c*, in month *m* of year *y*;
$SPEI_{c,y,m}$, the moving averages of Standardized Precipitation Evapotranspiration Index (SPEI) of the current and previous 11 months in county *c*, in month *m* of year *y*;
$nonsmokePM_{c,y,m}$ (unit: $\mu g/m^3$), the moving averages of non-wildfire smoke $PM_{2.5}$ concentration of the current and previous 11 months in county *c*, in month *m* of year *y*;
$smokePM_{c,y,m}$ (unit: $\mu g/m^3$), the moving averages of wildfire smoke $PM_{2.5}$ concentration of the current and previous 11 months in county *c*, in month *m* of year *y*;
$\beta_1$, the coefficient for non-wildfire smoke $PM_{2.5}$ exposure;
$\beta_2$, the coefficient for wildfire smoke $PM_{2.5}$ exposure;
$\beta_3$, the coefficient for SPEI;
$\alpha_c$, the fix effect term for county *c*;
$\alpha_{y,m}$, the combined fix-effect term for year *y* and month of the year *m*;
$\alpha_{s,y}$, the combined fix-effect term for state *s* and year *y*;
$\alpha_{s,m}$, the combined fix-effect term for state *s* and month of the year *m*;
$Pop_{c,y}$, the number of populations in county *c* in year *y*, included as an offset.

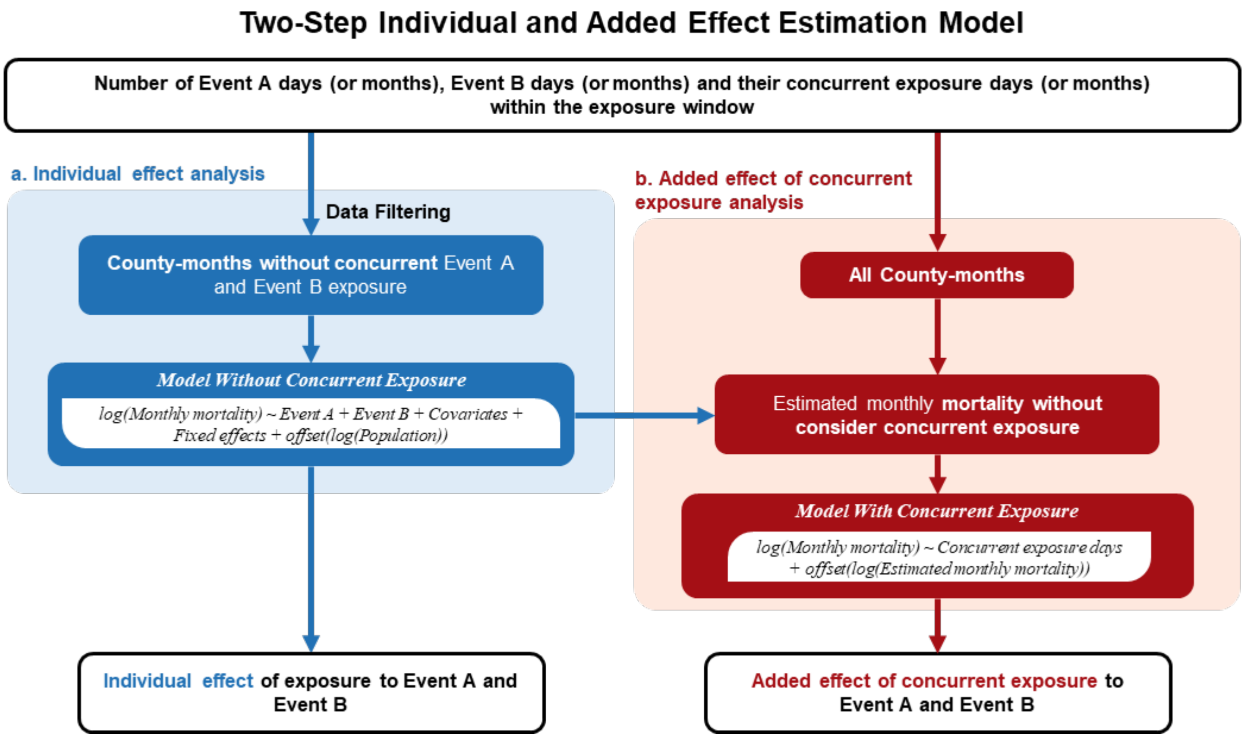

**Figure S1 Framework of the Two-Step Individual and Added Effect Estimation (TIAE) Model.**

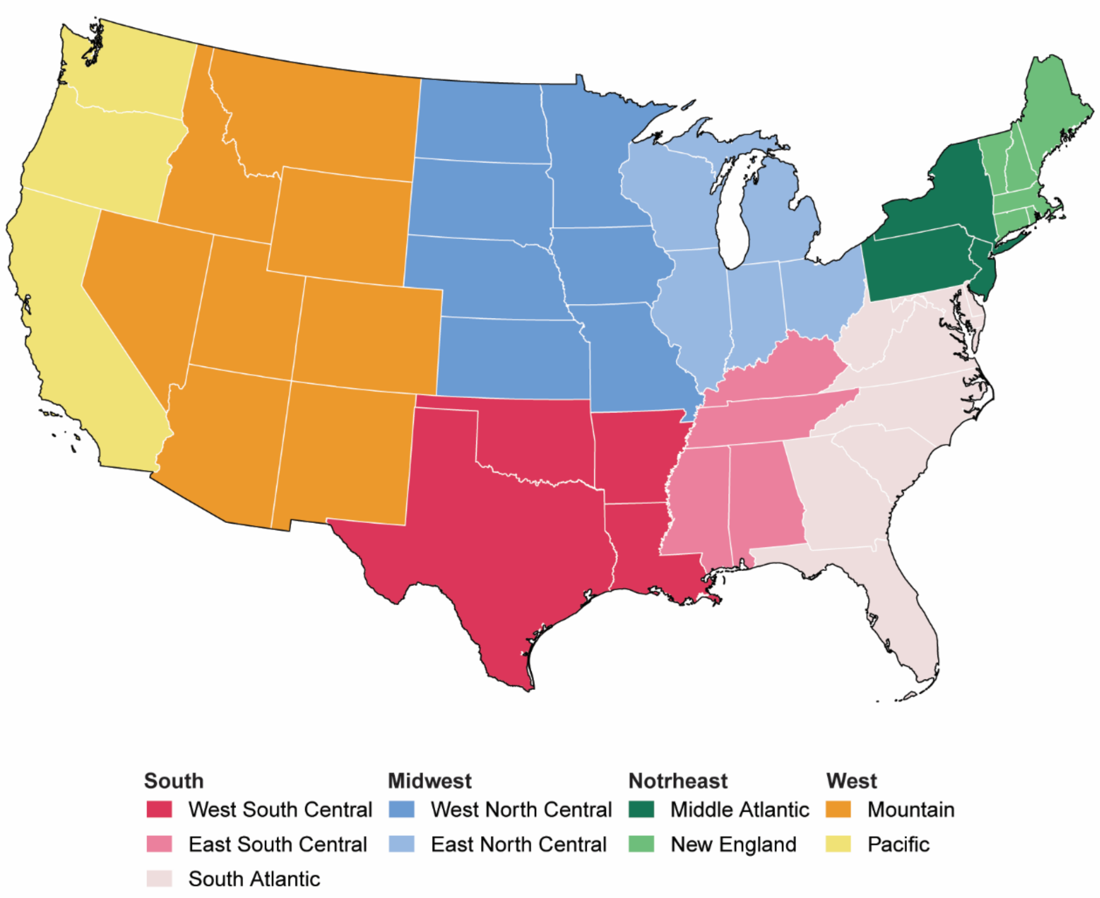

**Figure S2 Census Bureau-designated regions and divisions.**

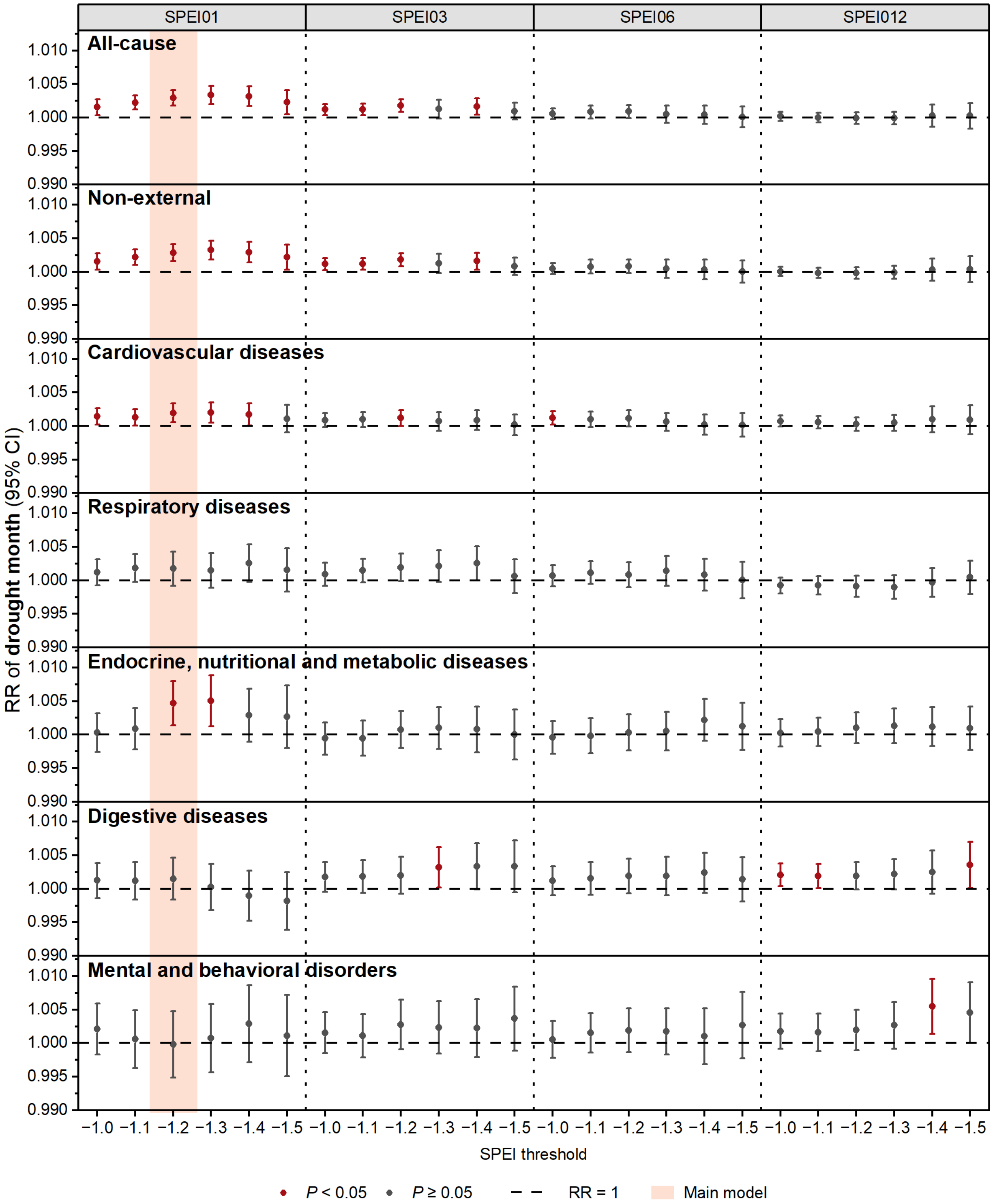

**Figure S3 Relative risk (RR and its 95% CI) of monthly cause-specific mortality associated with per additional month of drought during current and previous 11 months under different drought metrics.** Wildfire smoke day was identified as daily wildfire smoke $PM_{2.5} \geq 10\ \mu g/m^3$.

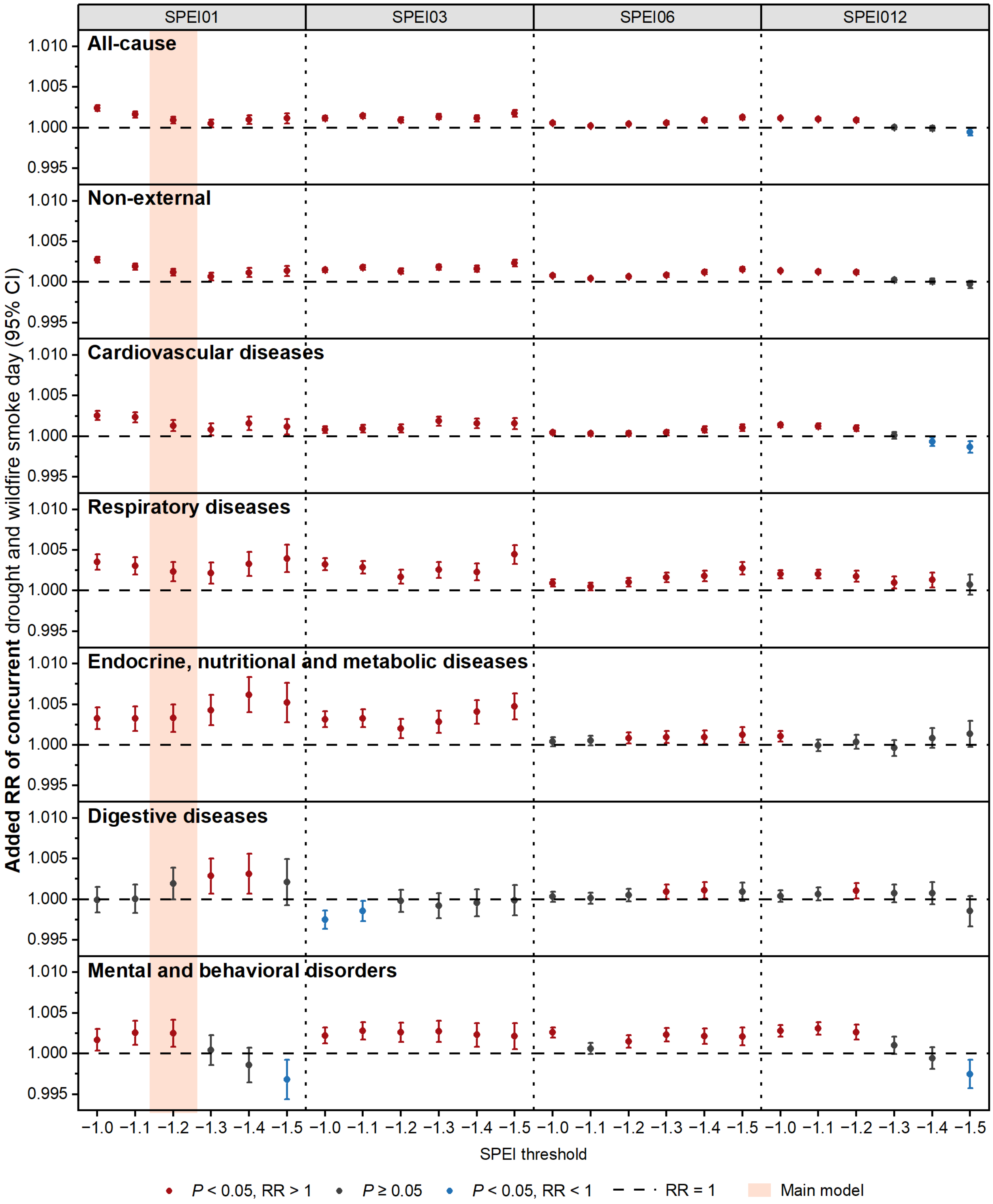

**Figure S4 Added relative risk (RR and its 95% CI) of monthly cause-specific mortality associated with per additional day of concurrent drought-wildfire smoke during current and previous 11 months under different drought metrics.** Wildfire smoke day was identified as daily wildfire smoke $PM_{2.5} \geq 10$ μg/m$^3$.

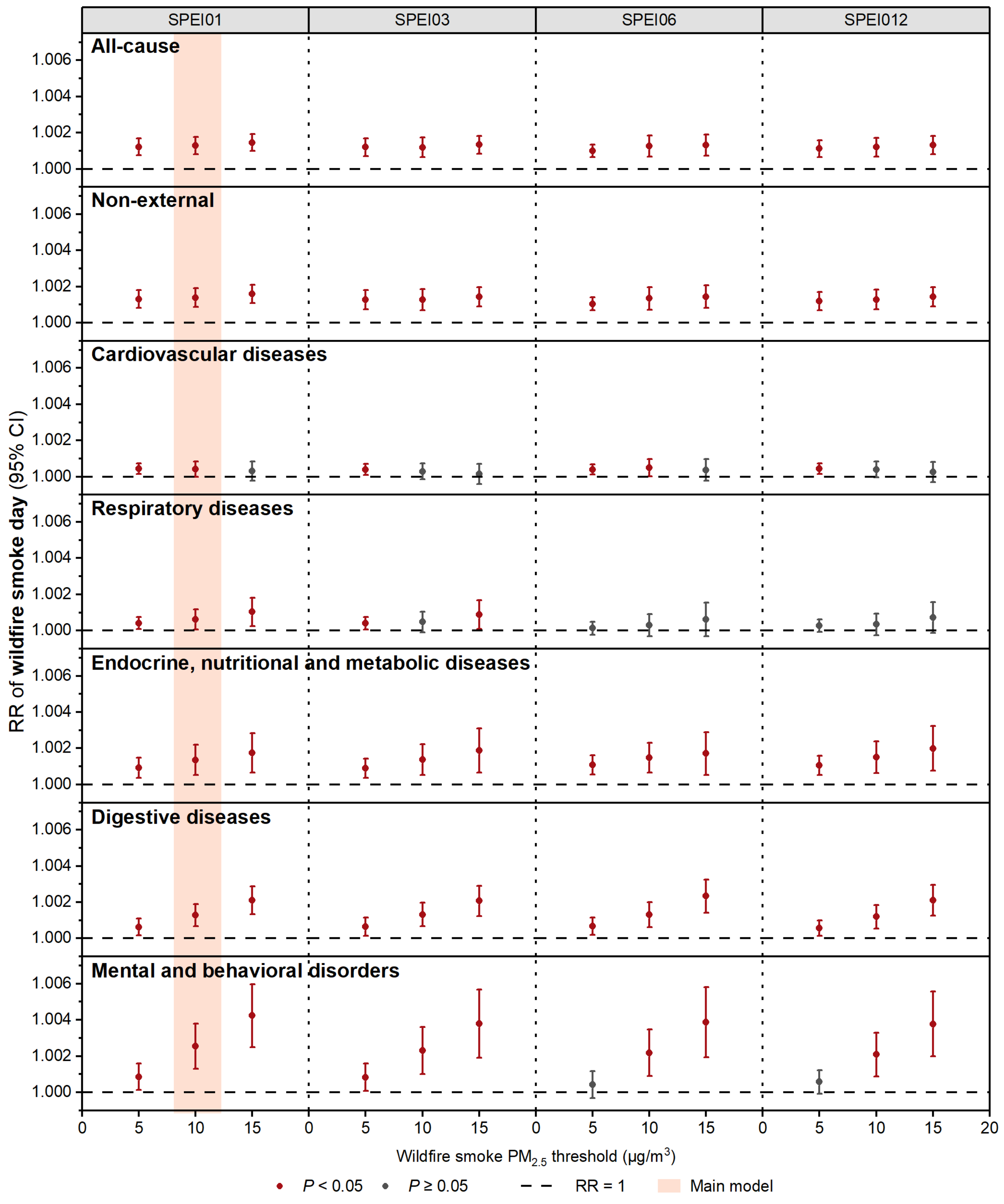

**Figure S5 Relative risk (RR and its 95% CI) of monthly cause-specific mortality associated with per additional day of wildfire smoke during current and previous 11 months under different wildfire smoke day metrics.** Drought day was identified as monthly SPEI < -1.2 under each SPEI timescale.

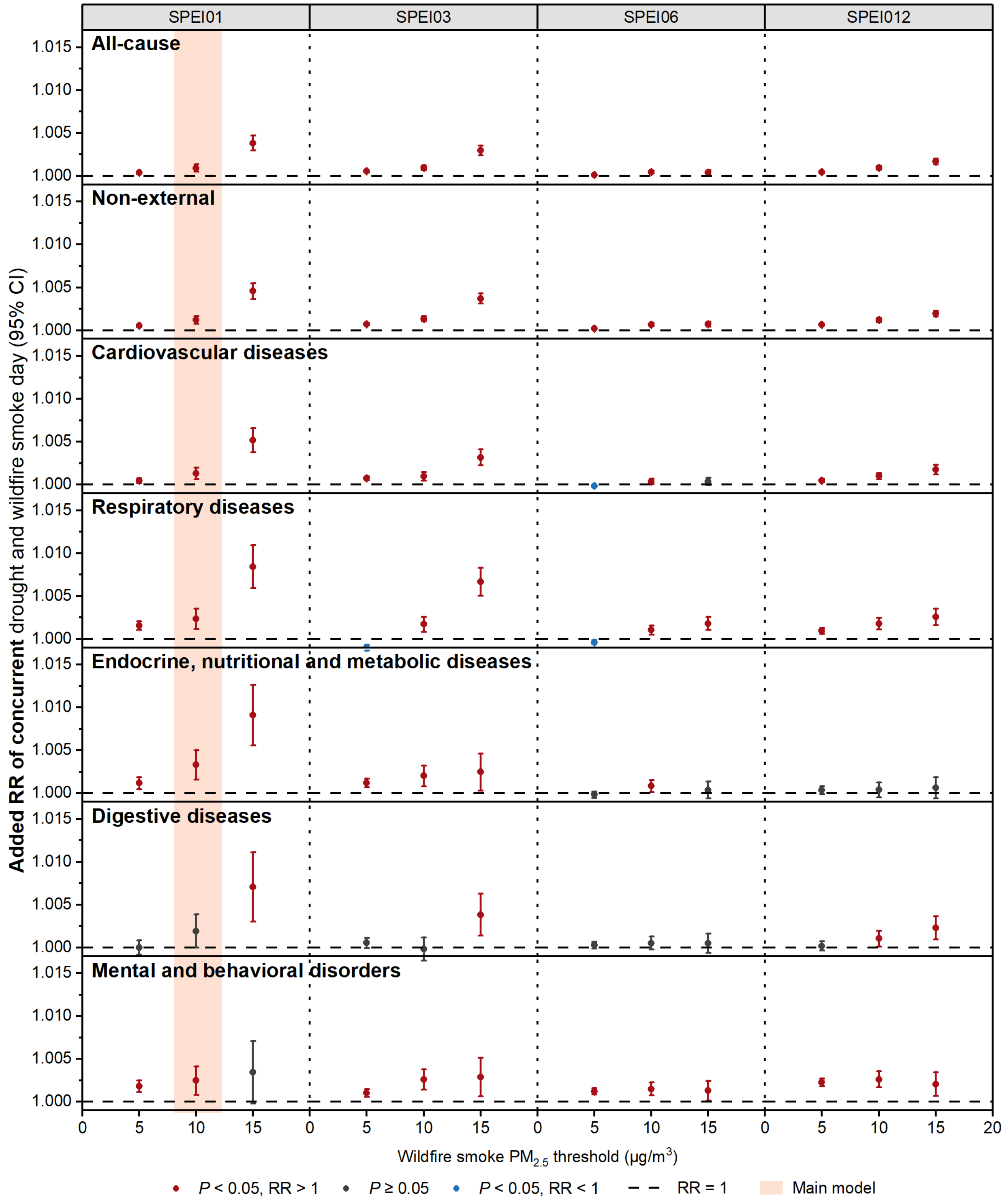

**Figure S6 Added relative risk (RR and its 95% CI) of monthly cause-specific mortality associated with per additional day of concurrent drought-wildfire smoke during current and previous 11 months under different wildfire smoke day metrics.** Drought day was identified as monthly SPEI < -1.2 under each SPEI timescale.

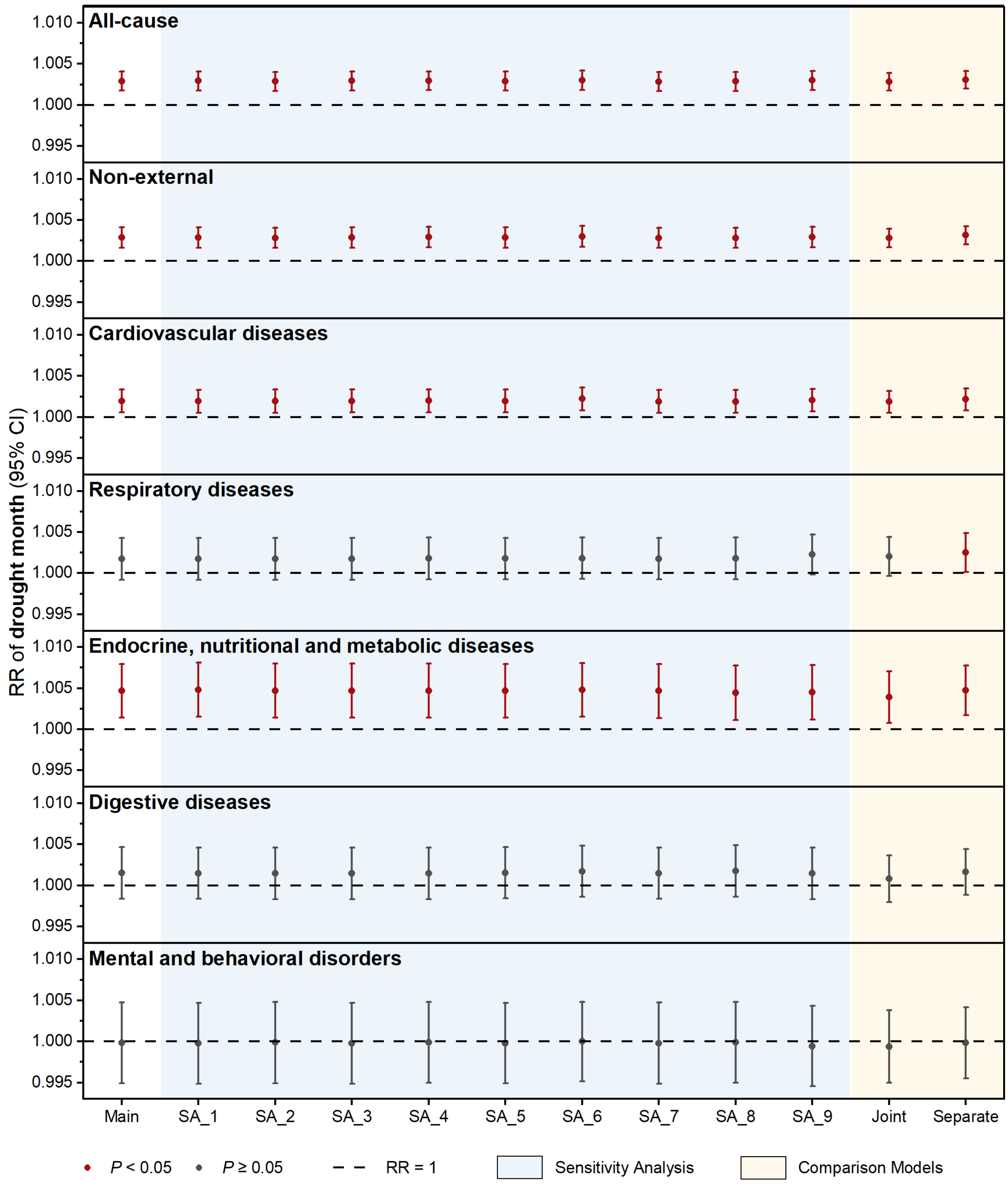

**Figure S7 Sensitivity analysis for relative risk (RR and its 95% CI) of monthly cause-specific mortality associated with per additional month of drought during current and previous 11 months using different models.** Model list was shown in Table S8.

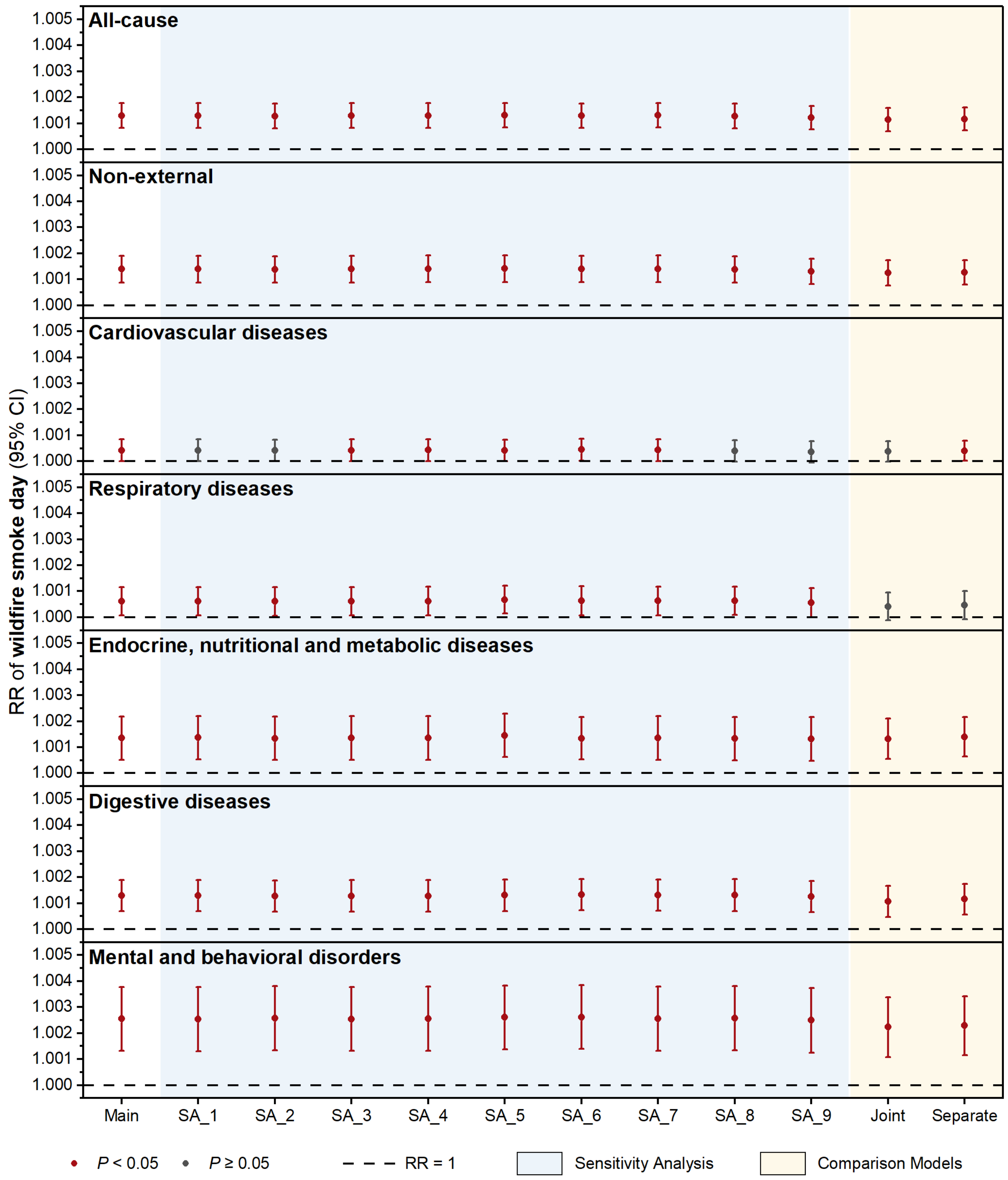

**Figure S8 Sensitivity analysis for relative risk (RR and its 95% CI) of monthly cause-specific mortality associated with per additional day of wildfire smoke during current and previous 11 months using different models.** Model list was shown in Table S8.

.

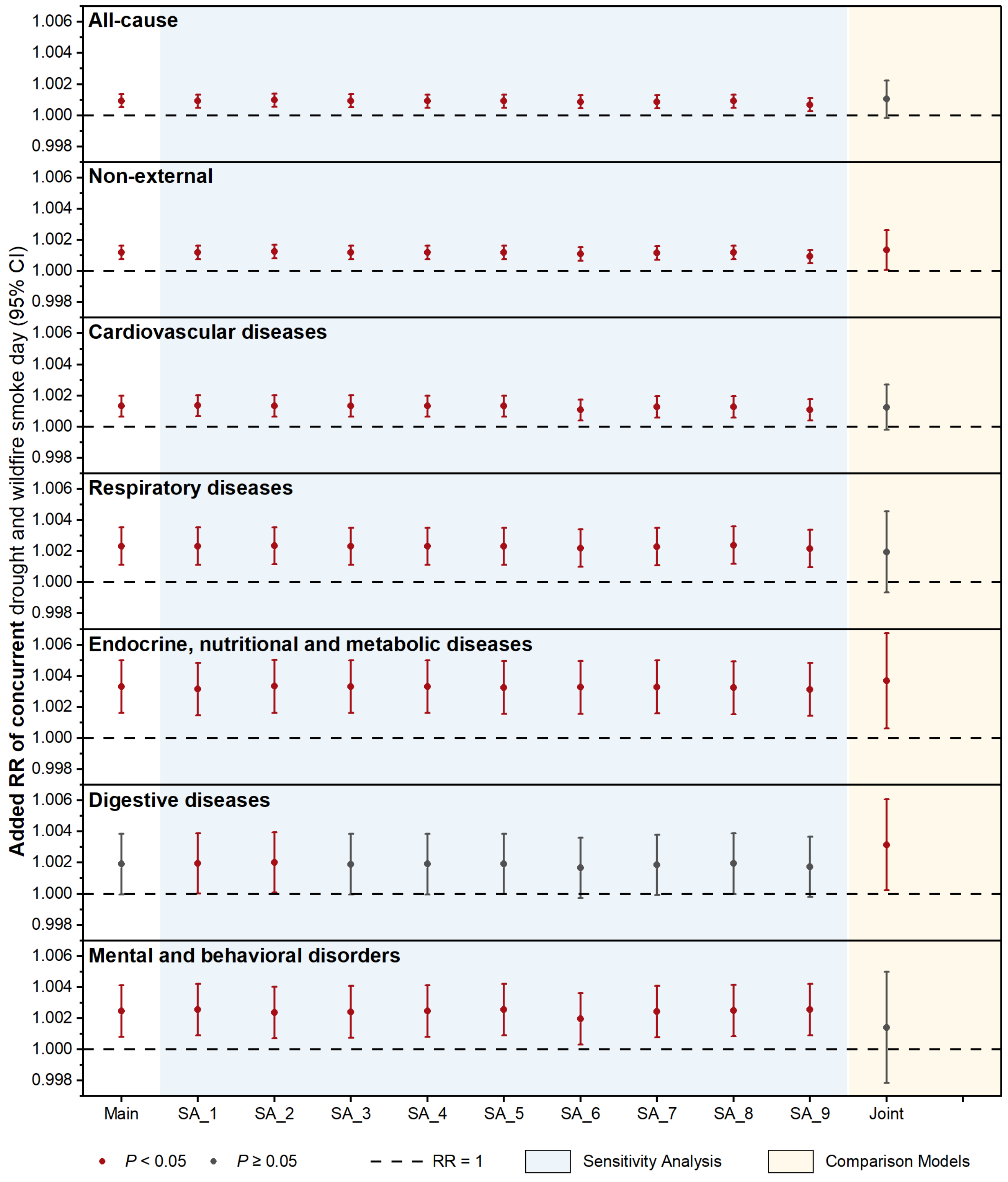

**Figure S9 Sensitivity analysis for added relative risk (RR and its 95% CI) of monthly cause-specific mortality associated with per additional day of concurrent drought-wildfire smoke during current and previous 11 months using different models.** Model list was shown in Table S8.

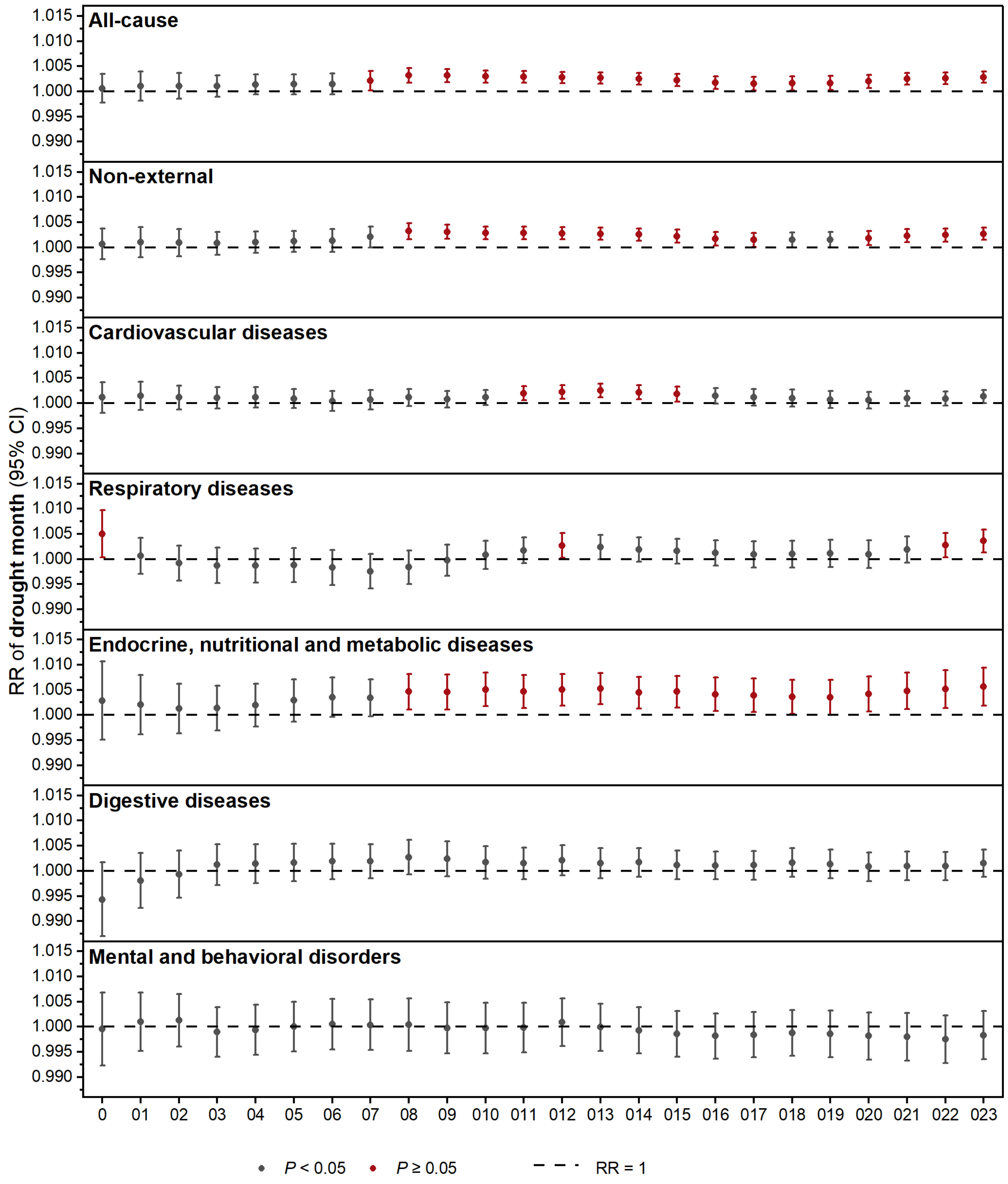

**Figure S10 Lag pattern test for relative risk (RR and its 95% CI) of monthly cause-specific mortality associated with per additional month of drought during current and previous 11 months.**

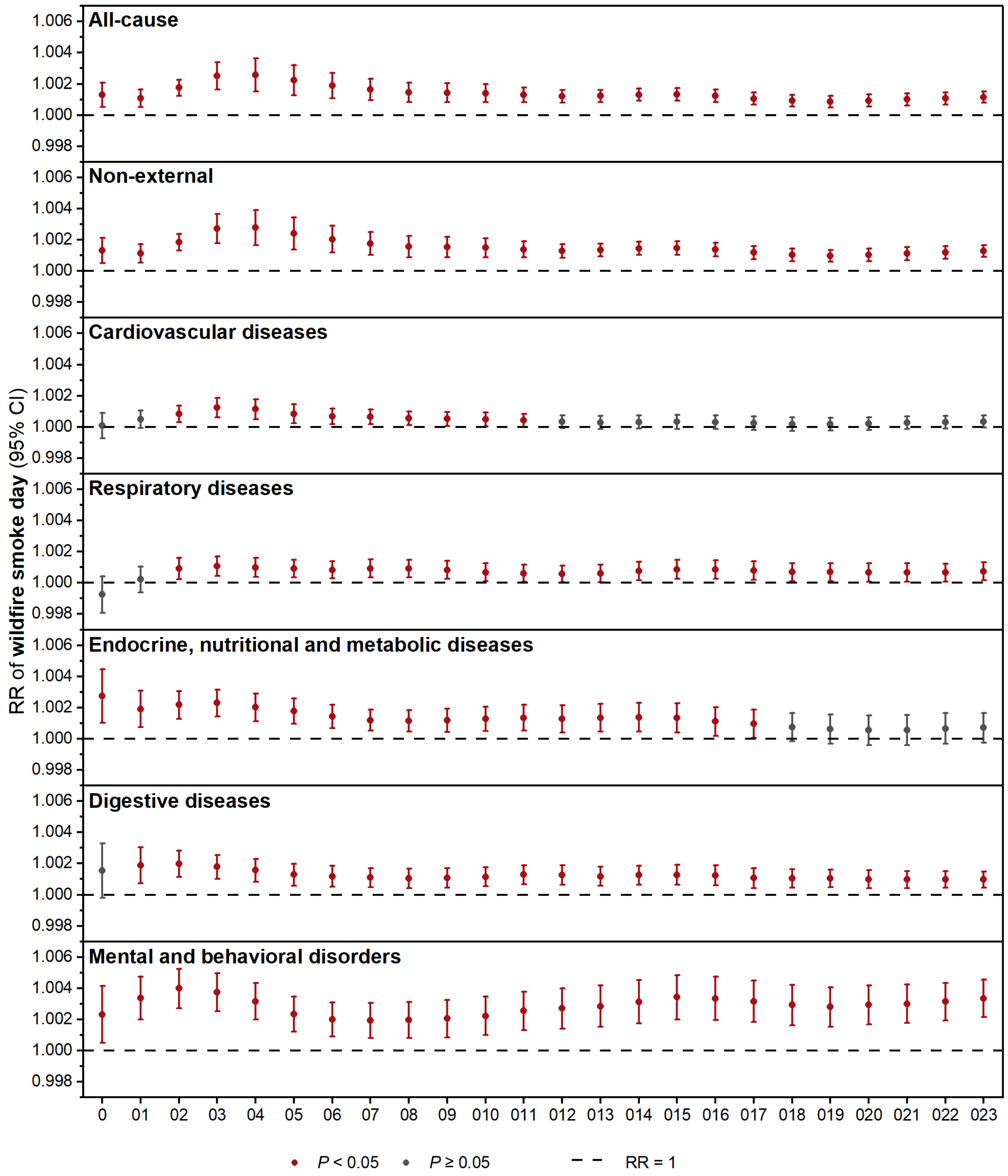

**Figure S11 Lag pattern test for relative risk (RR and its 95% CI) of monthly cause-specific mortality associated with per additional day of wildfire smoke during current and previous 11 months.**

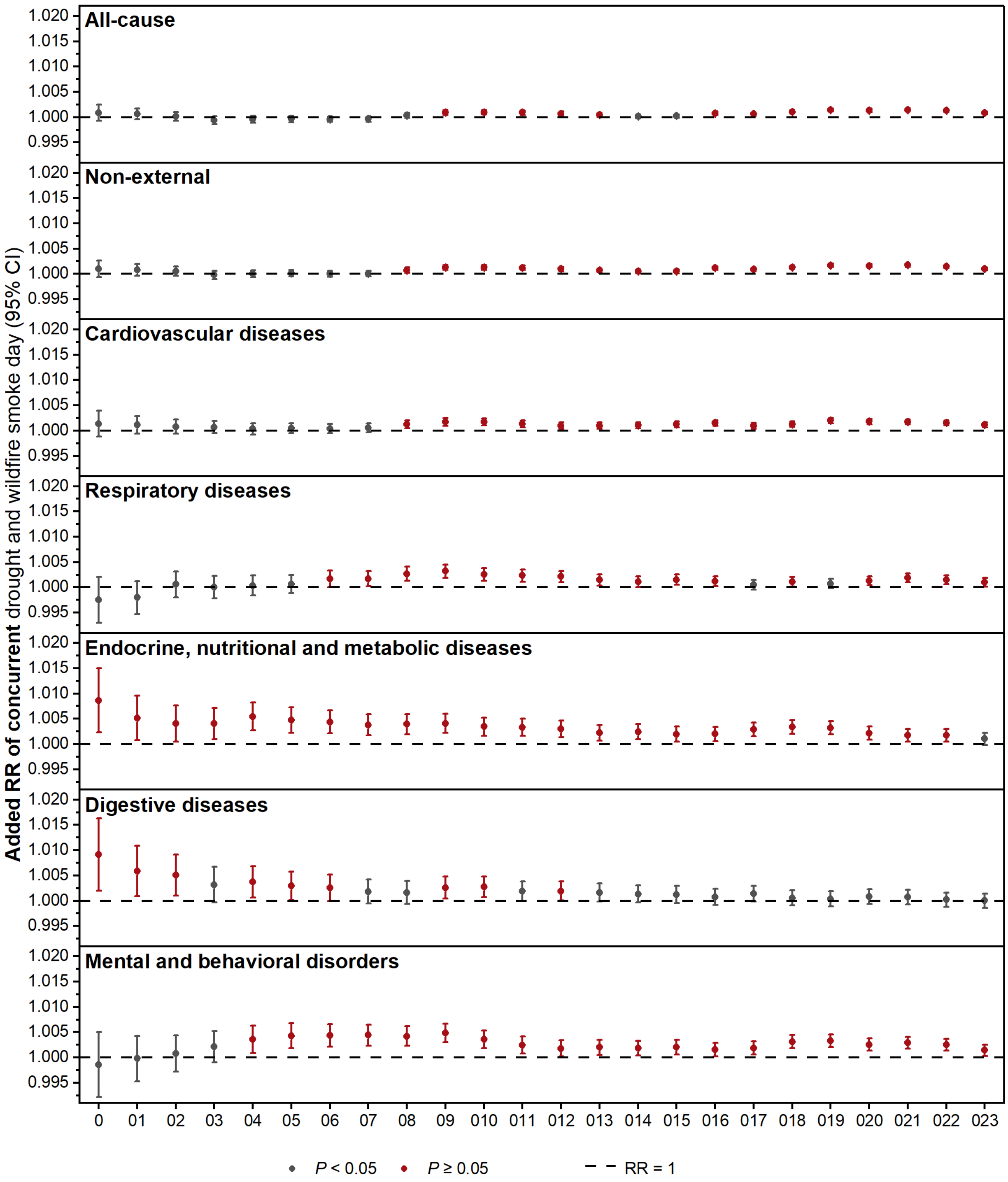

**Figure S12 Lag pattern test for added relative risk (RR and its 95% CI) of monthly cause-specific mortality associated with per additional day of concurrent drought and wildfire smoke during current and previous 11 months.**

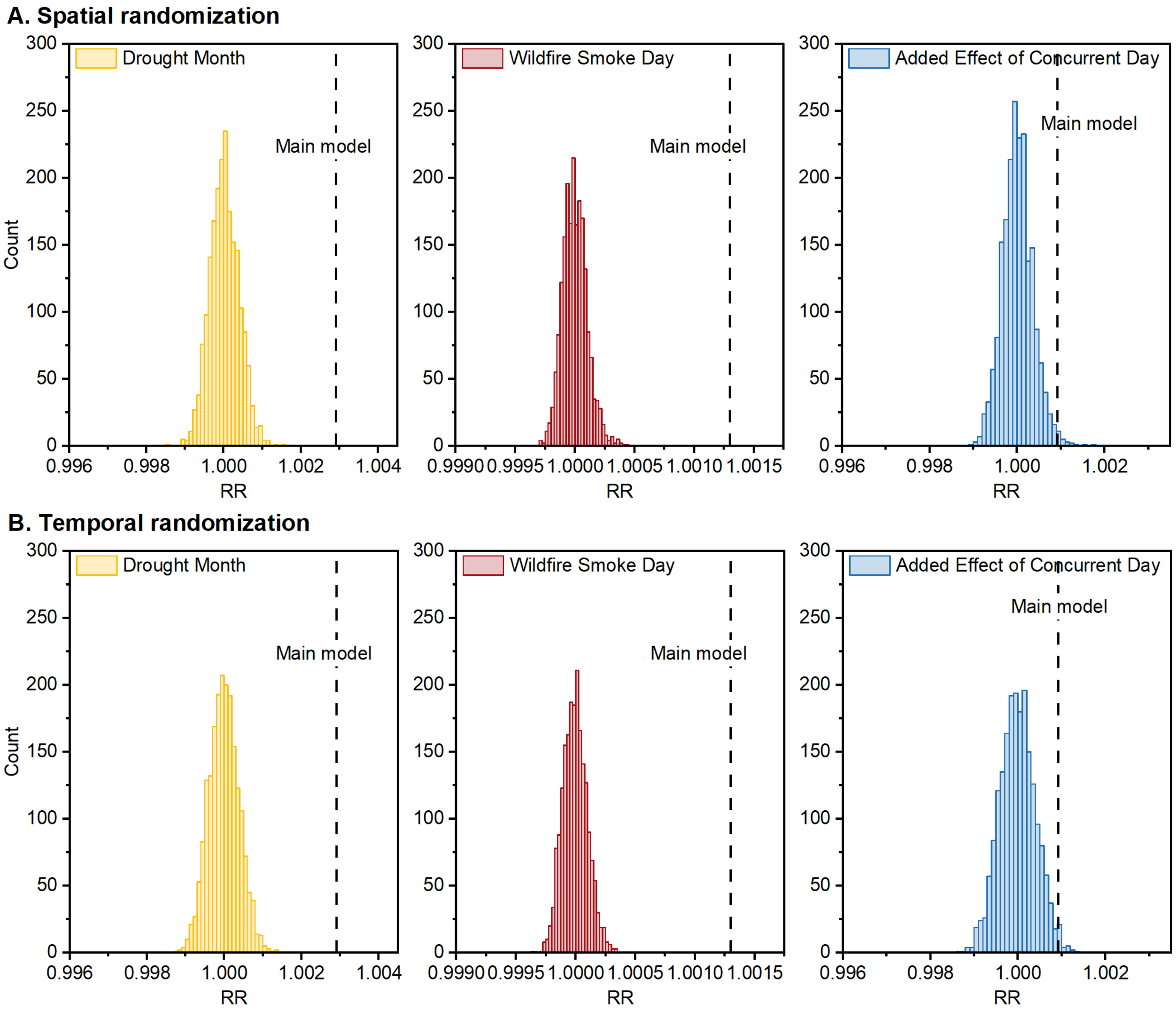

**Figure S13 Spatial and temporal randomization test for relative risk (RR and its 95% CI) of monthly all-cause mortality associated with drought and wildfire smoke, as well as added effect of their co-occurrence.**

For the spatial randomization test, we randomized county identifiers 2,000 times while holding the year and month constant. For the temporal randomization test, we randomized each year and month 2,000 times while holding county identifiers constant. The distributions of RRs from both the spatial and temporal randomization test were centered at one and our main estimations were on the upper side or outside the distributions, suggesting the model was not driven by spatial or temporal dependence due to misspecification.

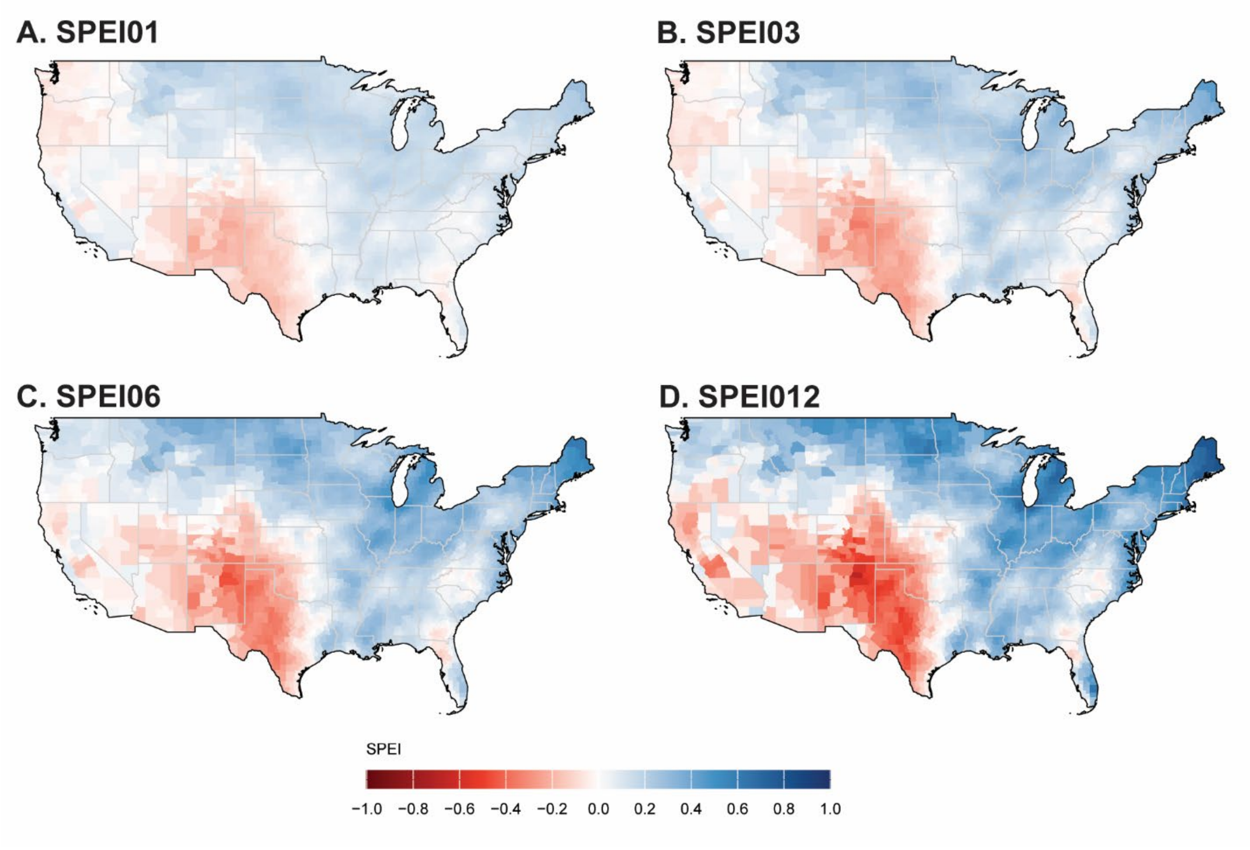

**Figure S14 Standardized Precipitation Evapotranspiration Index (SPEI) across 3,103 counties in the contiguous U.S. from 2006 to 2023 at timescales of 1, 3, 6, 12 months.**

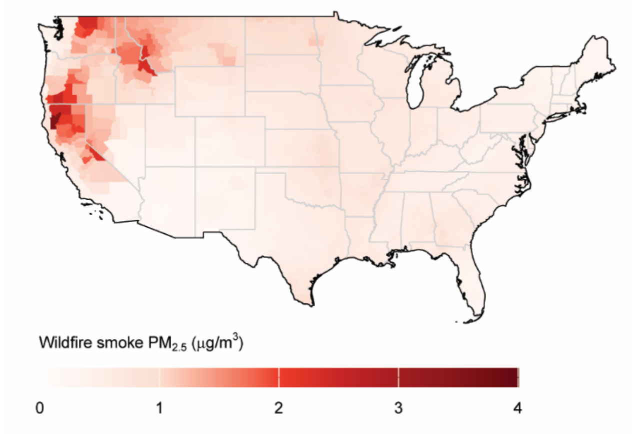

**Figure S15 Wildfire smoke $PM_{2.5}$ across 3,103 counties in the contiguous U.S. from 2006 to 2023.**
